# Caesarean section in Bangladesh, 2012-13 to 2025: facility birth, the private sector and small-area geography

**DOI:** 10.64898/2026.10.22.26366148

**Authors:** Fahima Sharmin Hossain, Kamrun Nahar, Fatematuz Zuhora, Umme Sultana Jesmin, Md Khairul Islam, Mohiuddin Sharif, Tanzin Naher, S. M. Ashraf Ul Hoque

**Affiliations:** Department of Obstetrics and gynaecology, Dhaka Medical College Hospital, Dhaka; Department of Obstetrics and gynaecology, Noakhali Medical college Hospital, Noakhali; Department of Obstetrics and gynaecology, Chhatak Upazila Health Complex, Sunamganj; Department of Obstetrics and gynaecology, Kurmitola Multicare Specialized Hospital, Dhaka; Department of Medicine, Dhaka Medical College Hospital, Dhaka; Department of Clinical Pathology, Dhaka Medical College Hospital, Dhaka; Department of Public Health, Independent University, Bangladesh (IUB), Dhaka

## Abstract

**Background:** In Bangladesh, caesarean section rose from one in five births in 2012-13 to one in two in 2025. We asked how much of this rise came from more facility births, a move to private facilities, and changing rates within sectors.

**Methods:** This repeated cross-sectional study used Multiple Indicator Cluster Surveys from 2012-13, 2019 and 2025 (26,539 recent mothers). A Kitagawa decomposition split the rise into parts. For 2025, we mapped caesarean use among facility births in 2,720 clusters and examined modelled travel time.

**Results:** The caesarean rate rose from 19.5% (95% CI 18.1 to 20.9) to 52.0% (50.7 to 53.2). More facility births made up 82.8% (78.9 to 87.7) of this 32.5 point rise and the move to private facilities 12.6% (10.1 to 15.0). In a one-step version, place of birth made up 94.7% (90.7 to 98.9). The private caesarean rate rose from 79.1% to 86.1%, while the public rate fell from 38.7% to 33.5%. Among private first births, which rarely follow a previous caesarean, the 2025 rate was 4.8 points (0.3 to 10.2) above the 2012-13 model’s prediction. The 2025 private rate was 85% to 87% in every wealth group. Longer travel time was linked with fewer caesareans, mainly through fewer facility births. Among private-sector births, travel time was not linked with caesarean (prevalence ratio 0.99 per 10 minutes, 0.97 to 1.01). Caesarean use among facility births clustered geographically; the private share of births in nearby clusters, not household factors, accounted for much of this.

**Conclusions:** In accounting terms, most of the rise came from births moving into facilities, mostly private ones, where caesarean was already common. The data cannot show how many caesareans were needed. If 2025 sector mix and rates persist, more facility births would keep raising the national rate.

## Introduction

Caesarean section rates are rising in every world region, reaching about 21% of births worldwide in 2010 to 2018, and are expected to keep rising until 2030 [1]. A caesarean saves lives when it is needed [2]. At the population level, rates above about 10% are not associated with lower maternal or newborn mortality. This threshold is not a target for single facilities, since referral hospitals with high-risk patients may need much higher rates [2]. National rates rise for two reasons. More women give birth in health facilities, and facilities perform more caesareans per birth. From 2000 to 2015, the first reason accounted for about two thirds of the global rise [3]. More facility births reflect access, ability to pay, personal preference and clinical need [4]. More caesareans within facilities call for a review of why they are done [3,5].

Bangladesh shows this rise in an extreme form. Bangladesh Demographic and Health Surveys (BDHS) up to 2022 show caesarean use rising across districts and wealth groups, and private facilities performed about 85% of all caesareans [6,7]. In private facilities, the caesarean rate rose from about 55% in 2004-07 to 84% in 2018-22 [7]. Among all facility births, it rose from 58.5% in 2014 to 69.4% in 2022 [8]. Over the same years, the share of families who borrowed money or sold assets to pay for childbirth doubled [8]. Women who gave birth in private facilities were far more likely to have a caesarean in the 2014 BDHS and in the 2019 Multiple Indicator Cluster Survey (MICS) [9,10]. Other studies of the 2019 survey mapped planned (elective) caesareans by district [11]. At the same time, some rural and poor women still cannot get a caesarean they need [12].

Three questions remain open. First, earlier studies described the private sector’s role [6,7] or measured how social and economic factors shaped caesarean inequality at one point in time [13]. A global analysis split the rise into more facility births and more caesareans within facilities [3]. To our knowledge, no study has split the rise in Bangladesh this way, or separated a shift to private facilities from changes within each sector [6–8]. No study has tested whether rates within a sector changed once the changing mix of patients is taken into account. Second, we do not know whether the private-sector difference varies by wealth, or whether wealth mainly decides who reaches a private facility. Third, one study linked the 2017-18 BDHS to the 2017 Bangladesh Health Facility Survey and found hotspots in Rajshahi, Dhaka and Khulna divisions [14]. In that study, well-equipped facilities nearby raised the chance of a caesarean, more so for private facilities. However, it studied caesarean among all births, so it could not separate access to facilities from practice inside them.

We therefore aimed to explain the rise in caesarean section in Bangladesh between 2012-13 and 2025. We split the rise into three parts: more facility births, the move between public and private facilities, and changes in rates within each sector. We did this nationally and for each wealth group. We also tested whether rates within each sector stayed stable once we took the type of patient into account. Finally, using the 2025 cluster locations, we examined whether modelled travel time, household factors or local delivery-sector mix explained where facility births were most often caesarean.

## Materials and methods

### Study design and data

We analysed three nationally representative household surveys: the Bangladesh MICS of 2012-13, 2019 and 2025 [15–17]. The Bangladesh Bureau of Statistics ran each survey with technical support from the United Nations Children’s Fund (UNICEF). Each survey used a two-stage cluster sample. Survey areas were selected within strata, and households were then selected within each area. Trained interviewers administered the standard MICS household and women’s questionnaires. In 2012-13, fieldwork ran from December 2012 to April 2013 on paper questionnaires, and all forms were entered twice. In 2019 (January to June) and 2025 (February to June), interviewers used tablet computers. Supervisors monitored fieldwork daily, and in 2019 and 2025 one household per cluster was interviewed again as a check. We excluded MICS 2006 because it did not ask about caesarean delivery. We followed the Strengthening the Reporting of Observational Studies in Epidemiology (STROBE) guideline for cross-sectional studies [18] (S1 Checklist). The response rate for women aged 15 to 49 years was 86.9% in 2012-13, 93.7% in 2019 and 95.8% in 2025.

The 2025 survey also released the location of each survey cluster. To protect privacy, locations were moved at random within the subdistrict (upazila). Urban locations moved up to 2 km and rural locations up to 5 km, or 10 km for 1% of rural clusters [19]. The survey also released a file of cluster data. This file gives modelled travel time to the nearest health facility, taken from a 2019 global travel-time map [20]. The map includes all types of health facility, not only those offering obstetric care.

### Study population

We included women aged 15 to 49 years who had a live birth in the two years before the interview and a recorded place of birth (Table 1). We excluded 154 women. Place of birth was “other” or missing for 109, 14 and 23 women by round, and the caesarean answer was missing for 7, 1 and 0. The final sample was 7,750, 9,270 and 9,519 women (26,539 in total). Each woman contributed her most recent live birth, so estimates describe recent live births to surveyed women, not all births. Births fell in roughly the two years before fieldwork: late 2010 to 2013, 2017 to 2019, and 2023 to 2025 [15–17]. Earlier births in the window and stillbirths were not included. The eligibility rule was the same in all three rounds.

**Table 1.** Characteristics of women aged 15 to 49 years with a birth in the last two years, by survey round (weighted percent).

| Characteristic | 2012-13 | 2019 | 2025 |
| --- | --- | --- | --- |
| Women with place of delivery recorded, before exclusions, n | 7,866 | 9,285 | 9,542 |
| Analytic sample, n | 7,750 | 9,270 | 9,519 |
| Age 15 to 19 years | 11.7 | 13.6 | 14.4 |
| First birth (parity 1) | 36.6 | 34.7 | 38.0 |
| Education: none | 18.4 | 9.2 | 4.3 |
| Education: secondary or higher | 52.9 | 67.6 | 79.1 |
| Urban residence | 21.1 | 21.9 | 28.4 |
| Four or more antenatal care visits | 25.2 | 36.9 | 43.4 |
| Place of delivery: home | 67.2 | 46.4 | 28.6 |
| Place of delivery: public facility | 13.1 | 15.9 | 16.9 |
| Place of delivery: private facility | 17.9 | 36.0 | 52.8 |
| Place of delivery: NGO or unknown sector | 0.0 | 1.4 | 1.5 |
| Place of delivery: other or missing | 1.8 | 0.2 | 0.2 |

### Outcome and place of delivery

The outcome was birth by caesarean section. The survey asks about caesarean only after a facility birth, so we coded home births as not caesarean. No home birth had a caesarean recorded.

Public facilities were government hospitals, government clinics and health centres, community clinics and other public facilities. Private facilities were private hospitals, clinics and maternity homes. S1 Table (panel B) shows how every survey category was classed in each round. We combined non-governmental organisation (NGO) facilities and facilities of unknown sector, because there were too few to analyse alone. The 2012-13 survey had no NGO category. We checked our sector shares and caesarean rates against each survey’s published report (S1 Table).

### Covariates

We adjusted for the mother’s age group, number of births (parity: 1, 2, 3, 4 or more) and education (none, primary, secondary or higher). We also adjusted for household wealth quintile, urban or rural residence, division and four or more antenatal care visits. Women with no antenatal care were counted as having fewer than four visits. We treated “don’t know” answers as missing. Missing covariate data affected 0.5% or fewer women in each round (S1 Table, panel C), so we used complete cases and did not impute. Each survey built a wealth index by principal components analysis of household assets and dwelling characteristics, and quintiles rank households within each round [15–17]. We assigned divisions from district codes, using the 2025 boundaries for all rounds. Wealth, education and residence were measured at interview, up to two years after the birth. We selected these confounders a priori, based on known or plausible links with facility birth and caesarean delivery, not by statistical testing. S1 Fig shows the assumed selection into facility and sector of birth.

### Statistical analysis

All estimates used each survey’s sampling weights, strata and clusters. We used the survey package in R, with design-based (Taylor linearisation) variances [21]. Survey round entered models as a categorical term. Aim 1 was the main analysis, and its primary estimands were the decomposition components of the 2012-13 to 2025 change. Other results are secondary or exploratory, and Aim 2 is exploratory. We interpret results by effect size and interval rather than P values. We did not register a study protocol in advance. The decomposition, travel-time and clustering analyses were defined with the aims. The case-mix standardisation, first-birth analyses, E-values, scenarios, travel-time splines, placebo tests and spatial sensitivity analyses were added after internal and external review, and are post hoc.

#### Aim 1: splitting the rise

The national caesarean rate equals the share of births in facilities times the caesarean rate among facility births. The facility rate is an average of the sector rates, weighted by each sector’s share of facility births. We split the change between two rounds into parts that add up to the total, using the symmetric method of Kitagawa [22]. Following Boerma and colleagues [3], the first step separates more facility births from a change in the facility caesarean rate. In a second step, we split the change in the facility rate into a shift between sectors and changes in each sector’s rate. Formally, R = f c and c = Σ m_k_ c_k_. Here f is the facility share, m_k_ the share of facility births in sector k, and c_k_ its caesarean rate. Then ΔR = Δf c^-^ + f^-^ Δc, and Δc = Σ Δm_k_ c^-^_k_ + Σ m^-^_k_ Δc_k_. Bars are averages of the two rounds. We report the within-sector term for each sector as well as in total. We used this two-step version as the main analysis because it matches the global analysis [3] and isolates the shift between sectors. The split between the first two parts depends on the order of the steps. We therefore also report a one-step version over four places of birth: home, public, private, and NGO or unknown. We applied the method to each pair of rounds, to each wealth quintile and residence group, and to first births. The shares are accounting contributions under each decomposition, not causal fractions. We obtained 95% confidence intervals (CIs) from 1,000 Rao-Wu rescaling bootstrap replicates, which resample clusters within strata and rescale their weights. Every component was recomputed in each replicate, and we used percentile intervals.

For public and private births separately, we fitted survey-weighted Poisson models of caesarean on survey round, adjusted for all covariates. These models give prevalence ratios (PRs). We repeated the models for first births. A first live birth can follow an earlier caesarean only after a pregnancy loss, so this removes most repeat caesareans. For the private sector, we also used standardisation. We fitted a separate model of caesarean on all covariates for each round’s private births, so every coefficient could differ between rounds. We then applied each round’s model to the private patients of every round. This gives the rate expected if caesarean use, given measured characteristics, stayed as in one round while the mix of patients was that of another. We call the difference caesarean use not explained by measured case mix, meaning sociodemographic and reproductive characteristics, not clinical risk. It may reflect changes in provider practice, in unmeasured clinical risk or in women’s preferences. Bootstrap and prediction details are in S4 Table. Antenatal visits with private providers may themselves lead to planned caesareans. In a sensitivity analysis, we therefore left antenatal care out of these models. MICS does not record previous caesarean, which becomes more common as caesarean use rises. We therefore repeated the standardisation among first births, who are unlikely to have had one, and treat this as the main estimate. For the within-sector prevalence ratios, we calculated E-values [23]. An E-value is the smallest risk ratio that an unmeasured factor would need with both survey round and caesarean to explain away an estimate. Survey round marks time, not a treatment, so E-values describe how strong unmeasured confounding of the round contrast would need to be. They do not address selection into sectors. We also compared private with public births in each round and tabulated each sector’s rate by wealth quintile. Finally, we calculated two illustrative scenarios for 2025. The first raised facility birth to 90% with 2025 sector mix and rates. The second sent the extra facility births to public facilities at the 2025 public rate. Both hold sector rates fixed, and their intervals come from the same bootstrap. They are static accounting exercises, not forecasts.

#### Aim 2: access and geography in 2025

We related modelled motorised travel time to the nearest health facility to six outcomes along the path to care. These were facility birth, private-facility birth and caesarean among all women; caesarean among facility births; and caesarean among private and public births. Travel time is a general accessibility measure, because the nearest facility may not offer caesarean or emergency obstetric care. Travel time entered as bands (under 5, 5 to under 10, 10 to under 20, 20 to under 30, and 30 minutes or more) and per 10 minutes. We used survey-weighted Poisson models adjusted for age group, parity, wealth, residence and education, with and without division. The PR per 10 minutes is only a summary. We therefore also fitted restricted cubic splines with three knots, at the 10th, 50th and 90th percentiles (2.7, 12.2 and 27.3 minutes). We tested non-linearity with a Wald test of the non-linear term. We report crude and adjusted PRs. In sensitivity checks, we excluded clusters more than 60 minutes away and used walking time instead.

For geography, we grouped facility births by survey cluster and modelled the number of caesareans among them. We measured clustering on the map with Moran’s I, using residuals from binomial models, the eight nearest clusters and 999 permutations. Because caesarean is common among facility births, odds ratios from these models exaggerate the corresponding PRs; we use them to compare models, not as measures of size. We added groups of factors one at a time: household factors, travel time, neighbouring delivery-sector composition (the private share of facility births), and division. A final model added a smooth surface over the map (a two-dimensional thin-plate spline with basis dimension 80), fitted by restricted maximum likelihood [24]. A cluster’s own private share comes from the same births as its caesarean rate, and clusters had a median of two births. Our main measure was therefore the private share of facility births in the eight nearest clusters. Other measures used the private share in the rest of the district and the cluster’s own share. The eight nearest clusters lay a median of 7 km away (5.8 km urban, 7.3 km rural). We therefore also defined neighbours as all clusters within 10, 15 or 20 km. Any covariate averaged over neighbours is spatially smooth and may absorb clustering for that reason alone. We therefore ran placebo models with neighbours’ averages of elevation, 2020 rainfall and wealth quintile, and of the private share after random permutation across clusters. These main spatial models were unweighted. Survey weights are identical for all women in a cluster, so weighting changes only the relative influence of clusters. Sensitivity checks used quasi-binomial models of survey-weighted cluster rates, 4 and 12 neighbours, basis dimensions of 40 and 120, and leaving out one division at a time. None of these spatial models uses strata or clusters for variance, so spatial uncertainty is model based.

We mapped probabilities in the same way as published maps of child growth failure [25]. We fitted a binomial generalized additive model with a smooth spatial surface. From it, we drew 1,000 sets of coefficients from their approximate Bayesian posterior, a multivariate normal distribution using the smoothing-corrected covariance matrix. For each location, we calculated the probability that the caesarean rate among facility births was above the national rate. We also calculated the probability that it was above 80%. We repeated this after adjusting for household factors and travel time, and again after also adjusting for the nearby private share. We report locations with a posterior probability above 0.8 or below 0.2, and, as a stricter check, above 0.95 or below 0.05. These probabilities are conditional on the fitted model. They do not include survey-design variance, coordinate displacement or uncertainty about the choice of model. Map boundaries came from geoBoundaries [26].

We used R 4.6.1 with the survey (4.5), mgcv (1.9.4), spdep (1.4.2) and sf (1.1.3) packages. We repeated the main estimates, and fitted the travel-time splines, in Stata 17 using Stata 15 syntax. We did not adjust for multiple comparisons, because only the decomposition components were primary.

### Ethics

This study used anonymised survey data from the UNICEF MICS programme. The 2025 survey protocol was approved by the Ethical Review Committee of the International Centre for Diarrhoeal Disease Research, Bangladesh (icddr,b) (PR-24113) [17]. The 2019 protocol was approved by a government technical committee led by the Bangladesh Bureau of Statistics [16]. The 2012-13 report does not describe its ethical review [15]. The 2019 and 2025 surveys obtained verbal informed consent [16,17]. We used cluster locations under the MICS data-use terms, only at cluster level, and made no attempt to identify households. No further ethical approval was needed for this secondary analysis.

## Results

### Study population

Between 2012-13 and 2025, the share of mothers with secondary or higher education rose from 52.9% to 79.1% (Table 1). Home births fell from 67.2% to 28.6%. Births in private facilities rose from 17.9% to 52.8%, while births in public facilities changed little. Our place-of-birth shares matched the published reports to within 0.2 points (S1 Table). With the reports’ own denominators, our caesarean rates matched the published 19.1%, 36.0% and 51.8% exactly (S1 Table).

### Aim 1: decomposition of the caesarean rise

Within facilities, the private share of births rose from 57.6% to 74.2% (Table 2). In private facilities, the caesarean rate rose from 79.1% to 86.1%. In public facilities, it fell from 38.7% to 33.5%. After adjustment, private births were 1.87 times (95% CI 1.65 to 2.12) as likely as public births to be caesarean in 2012-13. By 2025 they were 2.46 times as likely (95% CI 2.27 to 2.67) (S2 Table).

**Table 2.**
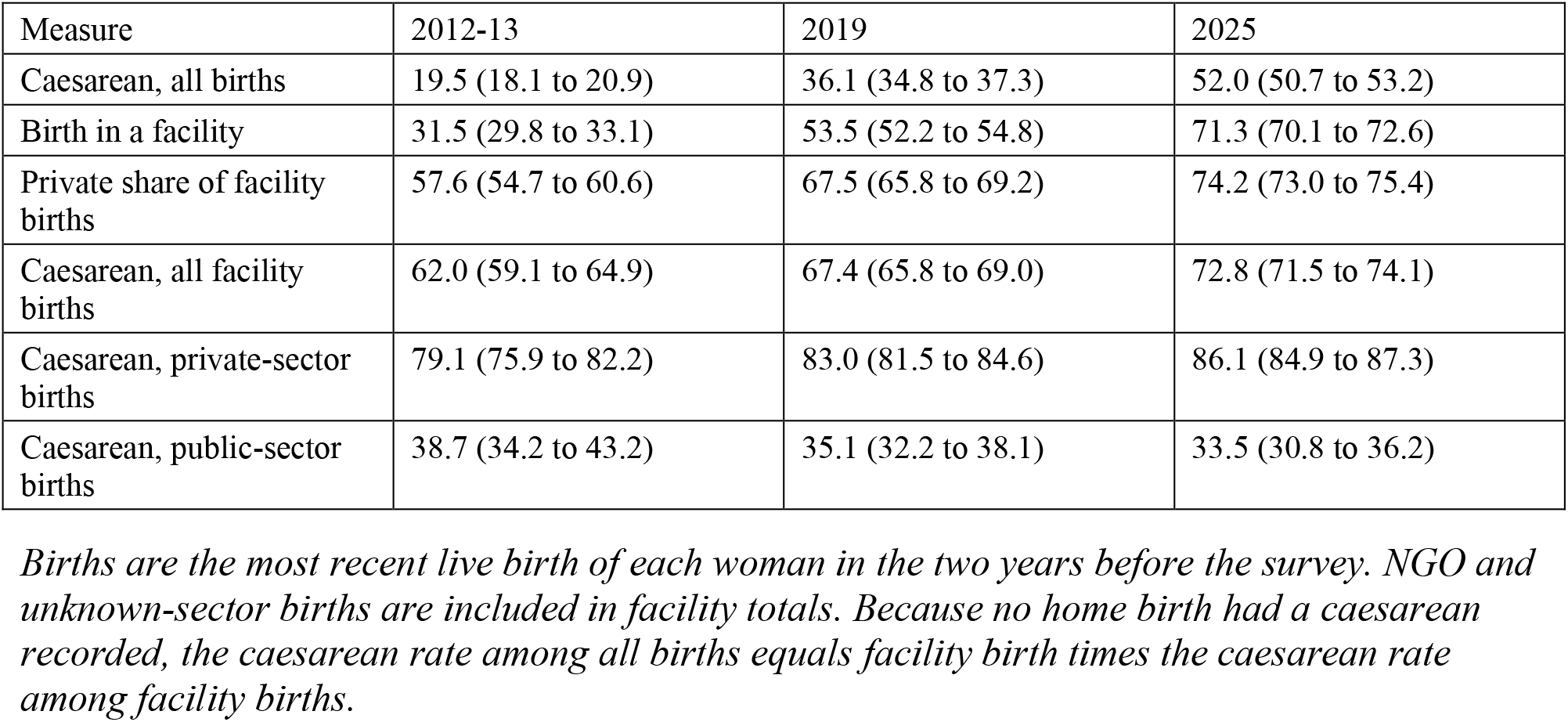
Facility birth, sector mix and caesarean rates by survey round (weighted percent, 95% CI).

The caesarean rate rose by 32.5 percentage points (30.6 to 34.4) between 2012-13 and 2025 (Fig 1A). More facility births accounted for 26.9 points, or 82.8% of the rise (78.9 to 87.7). The move from public to private facilities accounted for 4.1 points, or 12.6% (10.1 to 15.0). Changes in unadjusted rates within sectors accounted for 1.5 points (0.0 to 3.0), or 4.6% (−0.1 to 8.8). This small net term combined a rising private rate, which contributed 2.4 points (1.3 to 3.6), with a falling public rate (−0.9 points, −1.7 to 0.0) (S3 Table). In the one-step version, place of birth accounted for 94.7% (90.7 to 98.9) and rates within each place for 5.3% (1.1 to 9.3). In both versions, the within-sector part was larger from 2019 to 2025 than from 2012-13 to 2019 (S3 Table). With 2025 sector mix and rates, facility birth of 90% would give a national rate of 65.5% (64.4 to 66.8). If the extra facility births went to public facilities at their 2025 rate, it would be 58.2% (56.9 to 59.5) (S3 Table).

**Fig 1.**
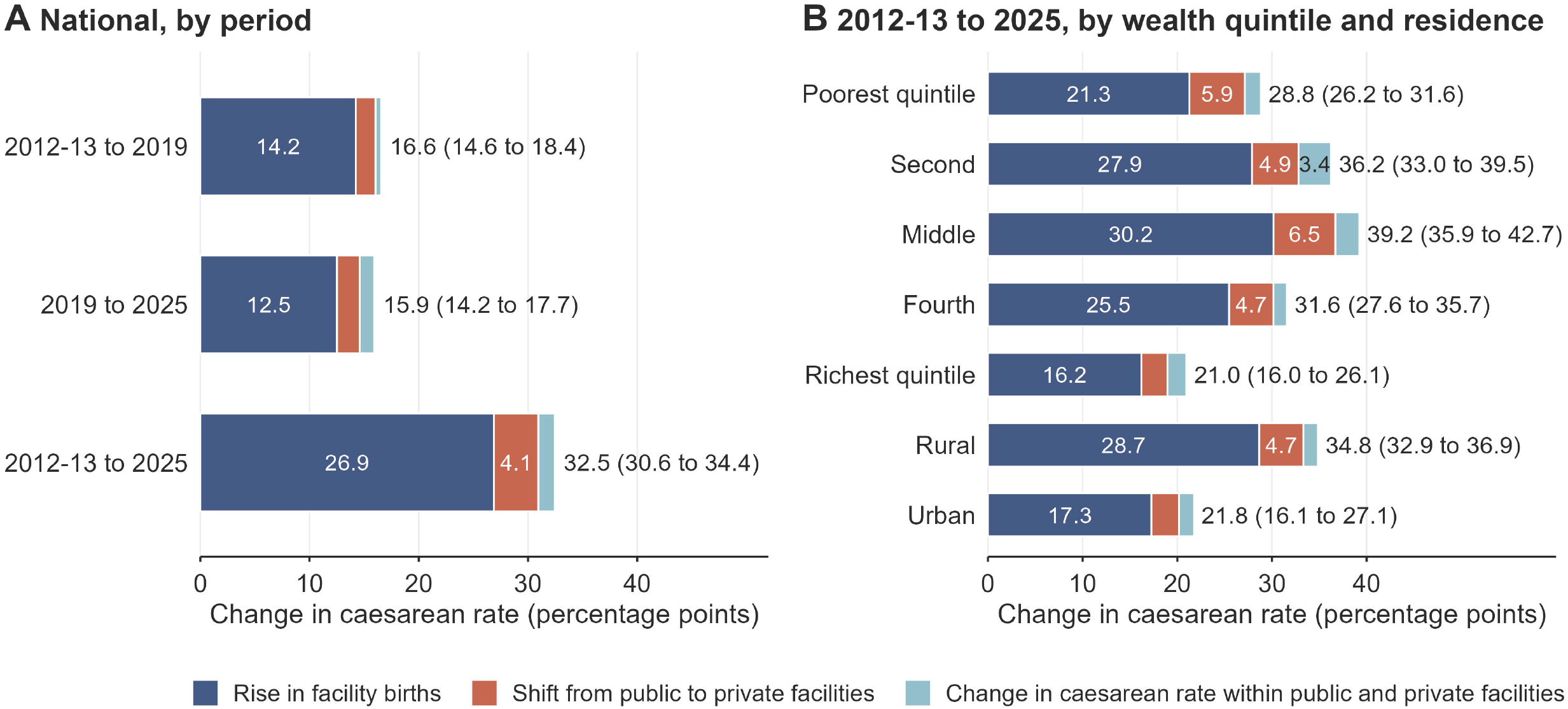
Decomposition of the rise in caesarean section, Bangladesh, 2012-13 to 2025. (A) Change in the national caesarean rate by period, split into the rise in facility births, the shift from public to private facilities, and the change in caesarean rate within public and private facilities. Values inside bars give components of 3 points or more; numbers at bar ends give the total change with 95% CI. (B) The same split for 2012-13 to 2025 within wealth quintiles and by residence. Design follows the facility and within-facility decomposition of Boerma and colleagues [3].

Among private patients, first births fell from 52.4% to 43.0%, and women from the two poorest wealth quintiles rose from 15.1% to 28.6% (S4 Table). Because quintiles rank households within each round, this is a change in relative wealth, not absolute wealth. Applying the 2012-13 private model to 2025 private patients predicted a rate of 76.8%. The observed rate was 86.1%, a gap of 9.3 percentage points (5.4 to 13.4). In the other direction, the 2025 model applied to 2012-13 private patients predicted 86.9%, against 79.0% observed. This gap was 7.9 points (4.3 to 11.7). The observed 79.0% differs slightly from 79.1% in Table 2 because this analysis excludes women with unknown antenatal care. Leaving antenatal care out of the models gave a gap of 9.5 points. After adjustment, the private caesarean rate in 2025 was 1.10 times (1.06 to 1.15) the 2012-13 rate. In public facilities the ratio was 0.94 (0.82 to 1.08) (S2 Table). The E-value for the private increase was 1.44 for the estimate and 1.30 for its lower limit (S2 Table).

Among first births, which exclude most repeat caesareans, the private rate rose from 81.7% to 86.1% (adjusted PR 1.06, 95% CI 1.00 to 1.12). The E-value was 1.32 for the estimate and 1.07 for its lower limit. The public rate for first births fell from 42.1% to 35.5% (S2 Table). In this group, changes in case mix barely moved the predicted rate. Applying the 2012-13 model to 2025 first births predicted 81.3%, against 86.1% observed, a gap of 4.8 points (0.3 to 10.2). In the other direction, the gap was 6.1 points (1.2 to 11.6) (S4 Table). Place of birth accounted for 97.4% of the rise among first births (S3 Table).

The rise was largest in the middle wealth quintiles and smallest in the richest (Fig 1B, S5 Table). In the poorest quintile, caesarean use rose from 5.5% to 34.4%. Over the same period, private-facility births in this group rose from 4.8% to 35.4% of all births. More facility births accounted for 73.8% of the poorest quintile’s rise, and the move to private facilities for 20.3% (15.4 to 25.3).

In private facilities, caesarean rates were similar across wealth quintiles (Fig 2, S6 Table). In 2025, the rate was 86.3% in the poorest quintile and 85.3% in the richest (unadjusted PR 0.99, 95% CI 0.95 to 1.03). The test of any wealth difference gave P = 0.90. In public facilities, the rate rose with wealth, from 21.2% to 49.7% (unadjusted PR 2.34, 95% CI 1.79 to 3.08). The wealth pattern differed between the two sectors (interaction P < 0.001). Adjusting for age, parity, education, residence and division gave similar results (S6 Table). Among all recent mothers in 2025, the share with a private caesarean was 30.6% in the poorest quintile and 59.4% in the richest. The PR was 1.94 (95% CI 1.78 to 2.13) (S6 Table).

**Fig 2.**
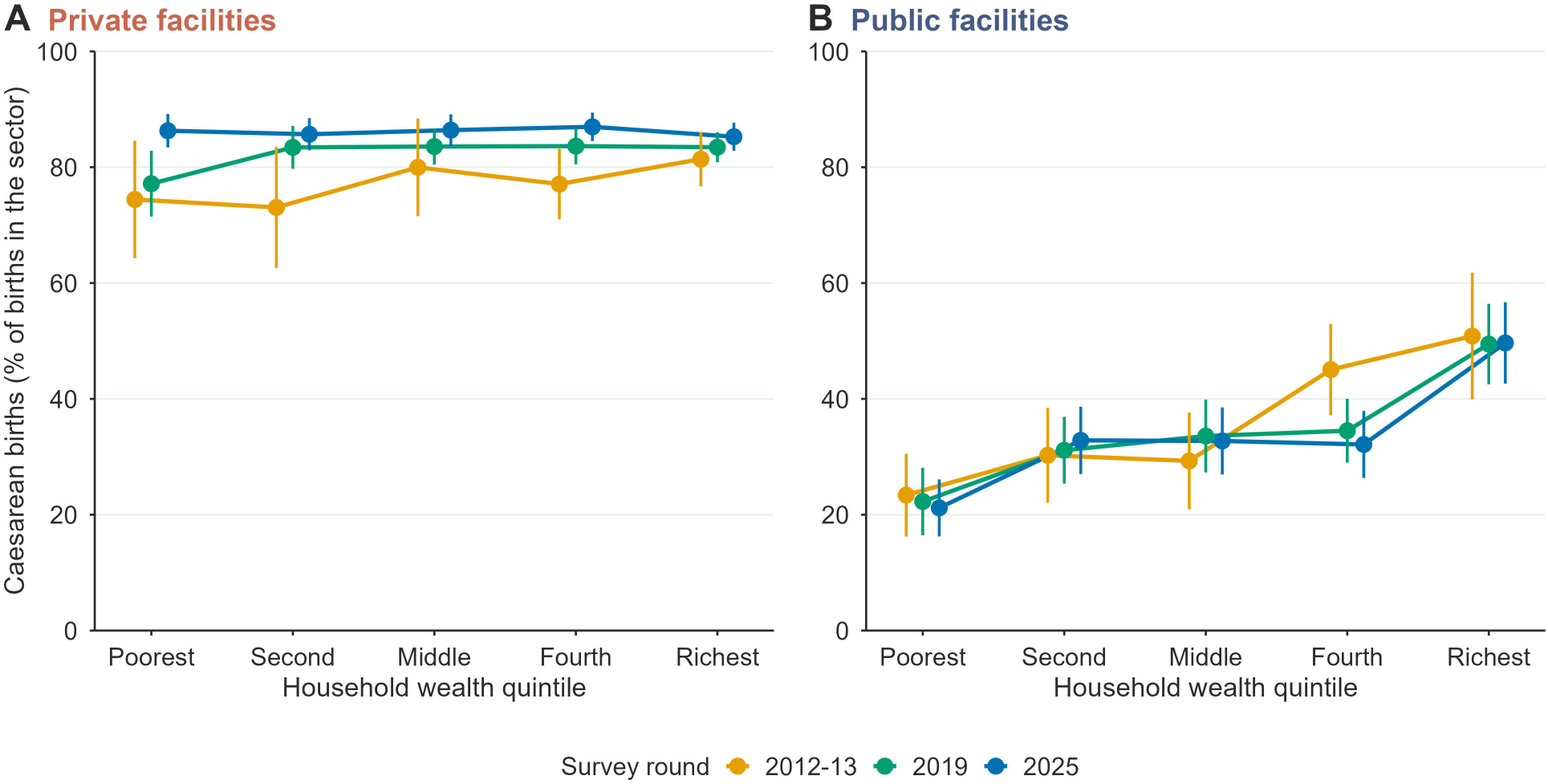
Caesarean rate by household wealth quintile within private and public facilities, by survey round. (A) Private facilities. (B) Public facilities. Points show weighted percentages with 95% confidence intervals; lines join the quintiles of each round. Values are in S6 Table.

### Aim 2: travel time and geography in 2025

The weighted median modelled motorised travel time to the nearest health facility was 11.0 minutes, and 6.0% of women lived 30 minutes or more away (S7 Table). Women who lived farther away were less likely to give birth in a facility, in a private facility, or by caesarean, to a similar degree (Table 3, Fig 3). Among facility births, the link with caesarean was weaker. After adjustment for division, it was no longer clear. Among private-sector births, travel time was not linked with caesarean. The private rate was 87.6% under 5 minutes and 83.7% at 30 minutes or more. Private-facility birth departed from a straight line (P for non-linearity < 0.001): PRs were slightly above 1 at 5 to 20 minutes and below 1 beyond 20 minutes. Facility birth showed no clear departure (P = 0.16), nor did caesarean among private births (P = 0.71) (S3 Fig). Public births within 5 minutes of a facility had a caesarean rate of 47.9%, against 25.6% to 31.1% farther away (P for non-linearity = 0.01). Results were similar when we excluded clusters more than 60 minutes away or used walking time (S7 Table).

**Fig 3.**
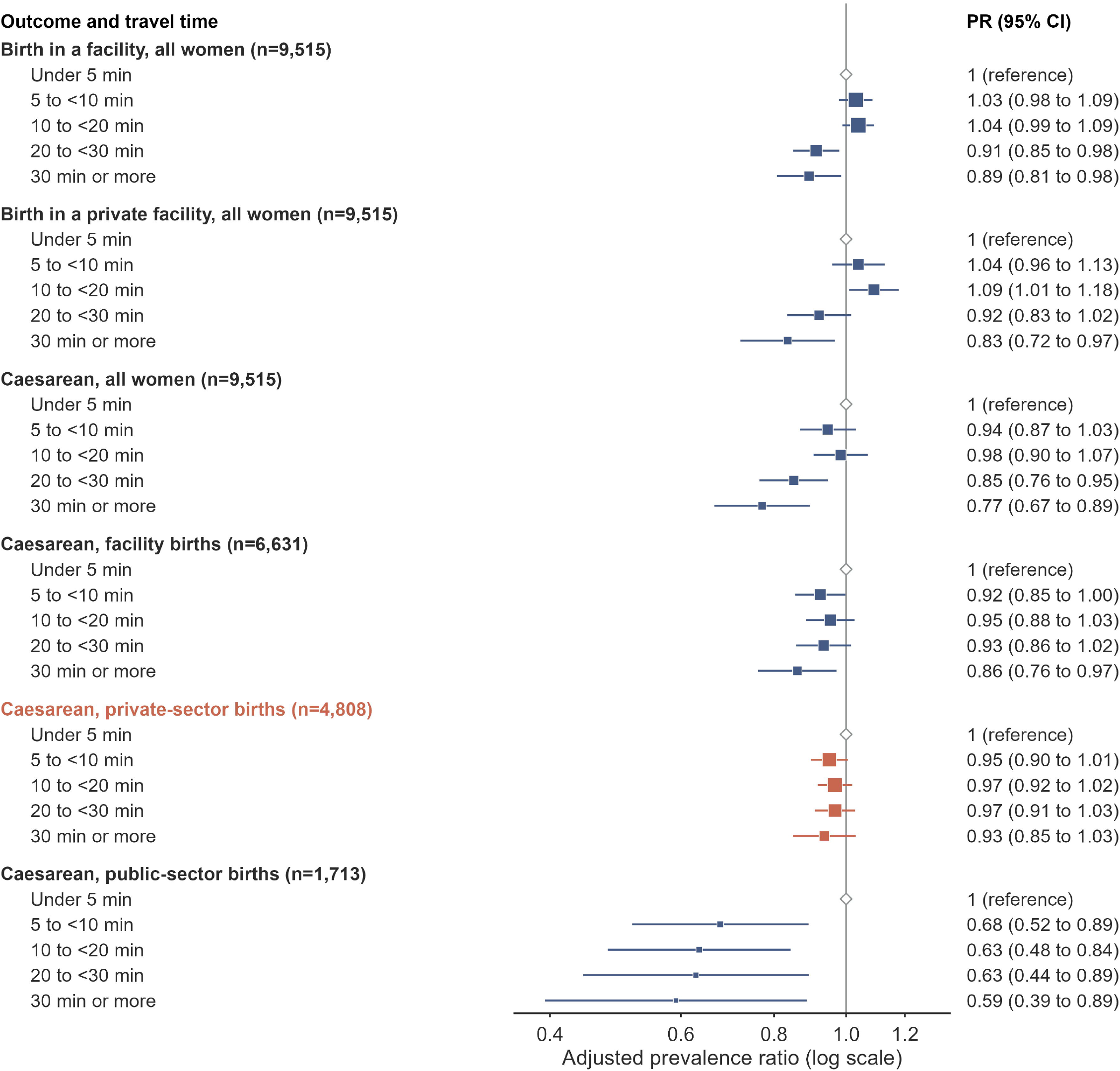
Modelled travel time to the nearest health facility and outcomes along the care pathway, 2025. Adjusted prevalence ratios with 95% confidence intervals for each travel-time band against under 5 minutes, with n for each outcome. Square area is proportional to the precision of the estimate. Models as in Table 3.

**Table 3.** Prevalence ratios per 10 minutes of modelled motorised travel time to the nearest health facility of any type, 2025.

| Outcome | Population | n | Crude PR (95% CI) | Adjusted PR (95% CI) | Also adjusted for division |
| --- | --- | --- | --- | --- | --- |
| Birth in a facility | All women | 9,515 | 0.92 (0.90 to 0.94) | 0.96 (0.94 to 0.98) | 0.96 (0.94 to 0.98) |
| Birth in a private facility | All women | 9,515 | 0.89 (0.87 to 0.92) | 0.94 (0.92 to 0.97) | 0.95 (0.92 to 0.98) |
| Caesarean | All women | 9,515 | 0.88 (0.85 to 0.90) | 0.93 (0.91 to 0.96) | 0.94 (0.91 to 0.97) |
| Caesarean | Facility births | 6,673 | 0.96 (0.94 to 0.98) | 0.97 (0.95 to 0.99) | 0.98 (0.96 to 1.00) |
| Caesarean | Private-sector births | 4,850 | 0.99 (0.98 to 1.01) | 0.99 (0.97 to 1.01) | 0.99 (0.97 to 1.01) |
| Caesarean | Public-sector births | 1,713 | 0.84 (0.76 to 0.92) | 0.91 (0.83 to 1.00) | 0.94 (0.86 to 1.02) |

*Survey-weighted Poisson models. Crude models include travel time only; adjusted models add age group, parity, wealth quintile, residence and education. Crude n is all 9,519 women, or all women in the subpopulation. All-women models include 9,515 of 9,519 women, because four lacked education data; facility-birth models include 6,673 of the 6,675 facility births for the same reason. The nearest facility may not offer caesarean or emergency obstetric care.*

The spatial analysis included 6,675 facility births in 2,720 clusters, a median of two per cluster (S8 Table). Caesarean use among facility births clustered on the map (Moran’s I 0.196, P = 0.001) (Table 4). Household factors did not reduce this clustering, and travel time reduced it only a little. Neighbouring delivery-sector composition accounted statistically for much of it: the private share of births in nearby clusters reduced Moran’s I to 0.070. Placebo averages over the same neighbours reduced it far less: to 0.176 for elevation, 0.155 for rainfall and 0.172 for wealth. A permuted private share left it at 0.183 (S8 Table). Each 10-point higher private share was linked to 1.30 times the odds of caesarean (1.26 to 1.35) (S8 Table). The private share in the rest of the district gave a similar result (Table 4). Division and a smooth spatial surface removed the remaining clustering. There was little extra variation (Pearson dispersion 1.07; S8 Table).

**Table 4.** Spatial clustering of caesarean among facility births, 2025 (2,720 clusters).

| Cluster-level model | Moran's I | Permutation P | Deviance explained (%) |
| --- | --- | --- | --- |
| No covariates | 0.196 | 0.001 | 0.0 |
| Household factors | 0.199 | 0.001 | 3.3 |
| Plus travel time | 0.183 | 0.001 | 4.2 |
| Plus neighbouring private share | 0.070 | 0.001 | 10.0 |
| Plus division | 0.036 | 0.001 | 15.0 |
| Plus two-dimensional spatial smooth | 0.009 | 0.142 | 20.0 |
| Alternative: district private share, own cluster excluded | 0.070 | 0.001 | 13.5 |
| Alternative: own-cluster private share | 0.130 | 0.001 | 27.1 |

*Moran's I of Pearson residuals from binomial models of each cluster's caesareans among its facility births, with eight nearest neighbours and row-standardised weights. Pearson residuals divide observed minus expected caesareans by the binomial standard deviation, so clusters of different size are on one scale. The permutation test reassigns residuals across cluster locations 999 times. Household factors: mean wealth quintile, urban residence, mean age group, and shares of first births, secondary education and four or more antenatal visits. The last two rows replace the neighbours' share in the travel-time model. The own-cluster share is built from the same births as the outcome, so it explains more deviance mechanically. Its sampling noise is not spatially structured, so it removes less spatial clustering.*

Survey-weighted models showed the same pattern, as did models with 4 or 12 neighbours and other basis dimensions (S8 Table). We also defined neighbours as all clusters within 10, 15 or 20 km. The private share then reduced Moran’s I from 0.194 to 0.071, 0.179 to 0.058 and 0.166 to 0.041 (S8 Table). Leaving out Chattogram division reduced the clustering before adjustment to a Moran’s I of 0.079. In every leave-one-out model, the odds ratio per 10 points of private share ranged from 1.19 to 1.33 (S8 Table).

The posterior probability of exceeding the national rate of 72.8% was above 0.8 over a central and western belt (Fig 4B, S9 Table). This belt covered almost all of Khulna and Dhaka and much of Barishal and Rajshahi. The probability was below 0.2 over most of Sylhet and Chattogram. The probability of exceeding 80% was above 0.8 mainly in Khulna and Dhaka (S2 Fig). The pattern held with stricter thresholds of 0.95 and 0.05. With these thresholds, 99.5% of Khulna and 94.2% of Dhaka cluster locations stayed above 0.95, and 79.4% of Sylhet locations stayed below 0.05 (S9 Table). These are regional patterns of a smoothed surface, not separate estimates for each cluster. Clusters where the probability exceeded 0.8 had a caesarean rate of 81.9% and a nearby private share of 79.5%. Clusters where it was below 0.2 had a caesarean rate of 51.6% and a nearby private share of 58.5%. The two groups had similar average wealth, and the higher group was less urban (26.5% against 31.9%) (S9 Table).

**Fig 4.**
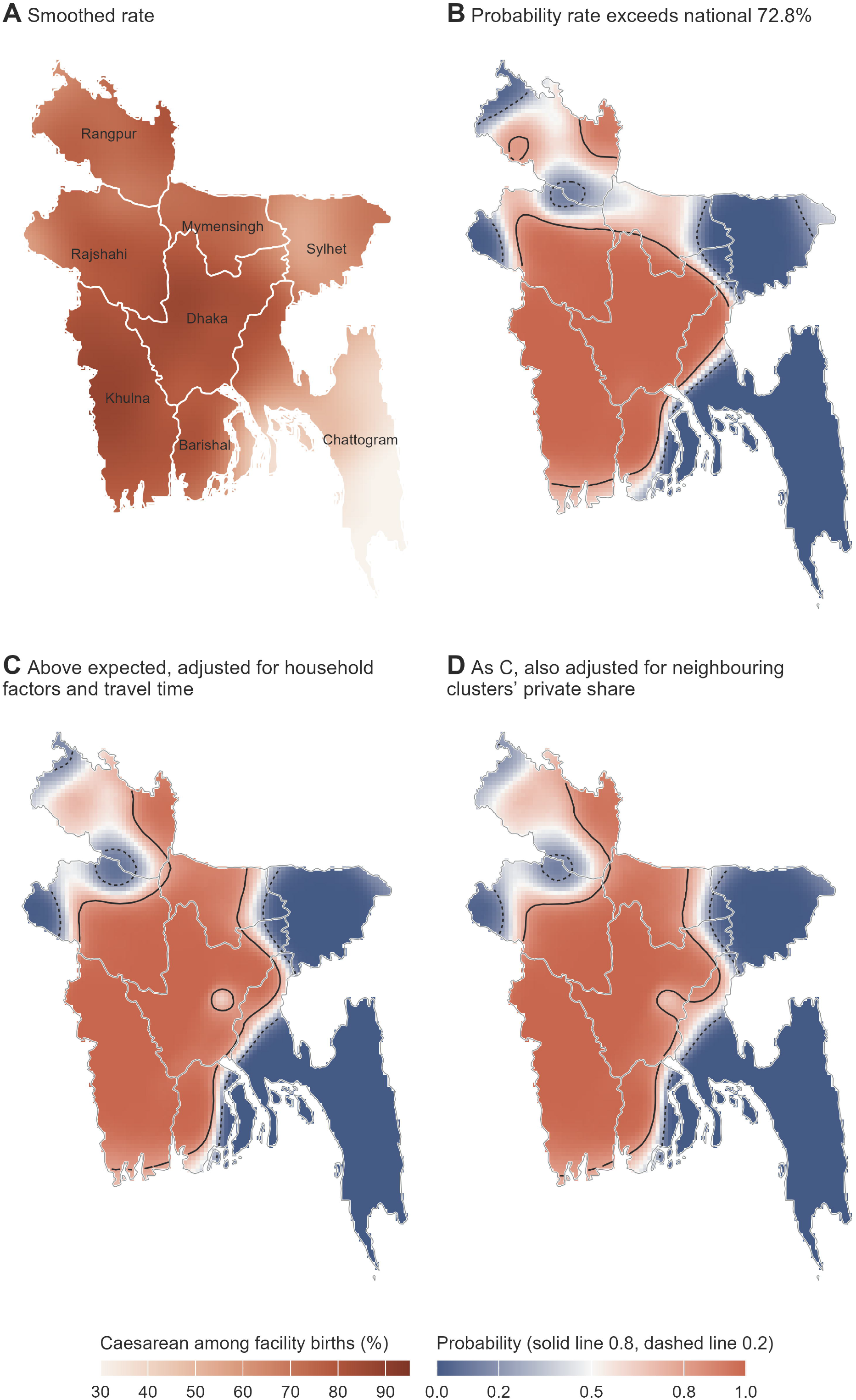
Caesarean section among facility births, 2025: probability maps. (A) Smoothed rate. (B) Probability that the rate exceeds the national rate of 72.8%. (C) Probability that the spatial effect is above zero after adjustment for household factors and travel time. (D) As C, also adjusted for the private share of facility births in the eight neighbouring clusters. Terracotta marks areas above, and navy areas below; solid lines mark a probability of 0.8 and dashed lines 0.2. Probabilities are conditional on the fitted model. Cluster coordinates are displaced for confidentiality. Boundaries: geoBoundaries.

Adjusting for household factors and travel time did not change this pattern (Fig 4C). Adjusting for the nearby private share also left the broad pattern in place (Fig 4D). The spread of the spatial effect also changed little. Its odds ratios ranged from 0.16 to 2.11 across the 5th to 95th centiles before this adjustment, and from 0.18 to 2.00 after it (S8 Table). Even within the private sector, the 2025 caesarean rate ranged from 73.5% in Chattogram to 92.4% in Khulna (S9 Table).

## Discussion

### Main findings

More facility births accounted for over four fifths of the caesarean rise in Bangladesh, and the move to private facilities for about one eighth. Private facilities already performed caesareans in four of five births in 2012-13. Their rate then rose further. Among first births, caesarean use not explained by measured case mix rose by about 5 to 6 points, with wide intervals. Within the private sector, caesarean use did not differ clearly by wealth or by modelled distance. Neighbouring delivery-sector composition accounted statistically for much of the small-area clustering, but not for a broad low-rate zone in the east.

### Sources of the rise

More facility births accounted for a larger share of the rise than the global two thirds [3], although that estimate covers other years and countries. The split between facility births and sector mix depends on the order of the steps, but the small net within-sector share does not. That net share hides a rising private rate partly offset by a falling public rate. Our rates agree with recent survey studies. In 2022, 69.4% of facility births were caesarean [8], and another analysis of the same survey reported 69% for hospital births [6]. The private-sector rate was 84% in 2018-22 [7]. These studies showed that the private sector dominates caesarean delivery. They did not separate the growth of the sector from changes in caesarean use within it [6,7]. Bangladesh is not unique. In India’s 2019-21 survey, place of delivery was the strongest predictor of caesarean [27]. In a national hospital-based cohort in Brazil in 2011-12, 87.5% of privately funded births were by caesarean [28].

Recent studies from other regions show similar sector gaps. In Arab countries of the Middle East and North Africa, caesarean use was higher in private facilities and in the richest wealth quintile [29]. In India, caesarean rates among first births fell slightly in public hospitals and rose in private hospitals between 2016 and 2021 [30]. Our first-birth results show the same divergence. In southern Europe, low-risk women had about twice the caesarean rate in private hospitals as in public ones [31]. In contrast with the Arab countries, the 2025 private rate in Bangladesh was similar across wealth groups.

Facility birth is a national goal with clear benefits. The problem is the caesarean rate a woman meets once she arrives. Our illustrative scenarios show that the national rate would rise further if new facility births followed the 2025 sector mix, and less if they went to public facilities. Both are static accounting scenarios, not forecasts. New facility users may differ from current ones, and sector rates may change; if private rates keep rising, both understate the national rate. Where new facility births happen matters as much as how many there are.

### Selection, case mix and repeat caesarean

Unadjusted rates alone do not show how caesarean use changed for similar women. Private patients became poorer and less often first-time mothers. Across all births, their rate rose 8 to 9 points beyond what this measured case mix predicted. But fewer first births means more women who may have had a previous caesarean. As caesarean use rose, this group almost certainly grew, so the all-births gap overstates any change. Among first births, the gap was 5 to 6 points, and both intervals only just excluded zero. The models lack presentation, multiple pregnancy, gestational age and complications, so the gap may reflect unmeasured clinical risk as well as provider practice or preference. The E-values point the same way. An unmeasured factor with a risk ratio of 1.32 with both round and caesarean could explain the first-birth increase. This is limited evidence of a modest rise in caesarean use not explained by measured case mix. It is not evidence of a large change in practice, nor evidence that practice stayed the same. In public facilities, the unadjusted rate fell, but after adjustment the change was close to none. The unadjusted fall may therefore mostly reflect a changing mix of patients as facility birth spread.

Repeat caesareans add further momentum. In settings with middle and low human development, women with a previous caesarean account for a growing share of caesareans [5]. MICS does not ask about previous caesareans. Among first births, the adjusted rise in private use was smaller than among all births. Part of the all-births rise may therefore reflect repeat caesareans. If first-birth rates stay high, more repeat caesareans are likely to follow in later pregnancies.

Selection into facilities and sectors also matters. Comparisons within sectors condition on facility birth, which wealth, travel time and unmeasured obstetric indication all affect. Among facility births, poorer women and women living farther away may more often have arrived with a complication. This can create or hide associations that do not exist among all women. Some women may also choose a private facility because a caesarean is already planned. Our method cannot separate births that moved to private care from women who moved there to have a caesarean.

### Wealth

Part of the poorest quintile’s rise came from the move into private facilities. This agrees with reports that caesarean use is spreading to poorer groups [7]. Wealth was linked to who reached a private facility, but not clearly to the caesarean rate once there. This flat gradient within private facilities may partly reflect selection, since poorer women in private care may be more clinically or financially selected than richer women. Among all mothers, however, the richest were about twice as likely as the poorest to have a private caesarean. India shows a similar pattern. There, the rich to poor ratio of caesarean was 4.0 in public facilities but only 1.4 in private ones [32]. In India too, social and economic factors mainly shaped access to facilities [27].

If selection is modest, the flat private gradient suggests that demand from wealthier women is not the main driver. It fits with practice norms set by private providers, with demand shared across all groups (such as fear of childbirth), or with both [33]. Poor women may also go to private facilities mainly when a caesarean is expected. Public hospitals receive referred complications, so the private against public ratio probably understates the private excess among women of similar risk. In public facilities, the wealth gap may reflect referral of complicated cases to large hospitals that richer women reach more easily. It may also reflect underuse of caesarean among poor women [12].

### Access and geography

Modelled travel time to the nearest health facility was linked mainly with whether and where women gave birth. It is a weak measure of access to caesarean care, because the nearest facility may not offer it. The lack of association within private births may therefore reflect a poor exposure measure. It does not show that access to surgical care is unimportant. This analysis is exploratory. The high public rate close to a facility probably reflects large district and specialist hospitals that receive referred complications. Shortages of anaesthetists and gynaecologists limit emergency obstetric care in many subdistrict hospitals [34].

Our maps agree with the 2017-18 study, which found hotspots in Rajshahi, Dhaka and Khulna linked to well-equipped private facilities nearby [14]. We add two findings. The pattern holds for caesarean among facility births, which separates practice from access, and household factors did not explain it. The low-rate zone in the east remained, even within the private sector, and Chattogram division accounted for much of the clustering. Moran’s I with eight neighbours measures fine-scale clustering among nearby clusters, while the smooth surface captures broad gradients. Neighbouring delivery-sector composition accounted for the fine-scale part, but not for the broad east-west gradient. Regional differences in medical practice, referral networks or demand may explain this, but our data cannot tell which.

Placebo covariates with similar spatial structure did not reproduce the drop in clustering seen with the nearby private share. That drop is therefore unlikely to be a smoothing artefact alone. Neighbouring delivery-sector composition shows where women gave birth, not facility supply, and neighbouring clusters need not use the same facilities. Its odds ratio describes an association with where neighbours gave birth, not the consequence of private supply. Linking the 2025 clusters to facility data, as was done for the 2017-18 survey [14], would test whether facility supply explains the pattern.

### Strengths and limitations

This study has several strengths. It uses three national surveys with the same questions, and our estimates match the published reports. Its methods account for the survey design. It reports two versions of the decomposition with CIs, and we repeated the key estimates in Stata. The 2025 survey adds many mapped clusters and modelled travel time.

The study also has limitations. First, MICS does not record why a caesarean was done. It also lacks the main clinical risk factors, such as previous caesarean, multiple pregnancy, the baby’s position, length of pregnancy, complications and referral. We therefore cannot say how many caesareans were unnecessary, and our standardisation covers only measured factors. Previous caesarean has almost certainly become more common, so the all-births standardised gap is likely to overestimate change in practice. The first-birth estimate avoids this but is less precise. Second, the decomposition shows where change happened, not what caused it. Third, analyses within sectors and the spatial analysis cover only facility births, so differences in who reaches a facility could create or hide associations. Fourth, the main spatial models were unweighted, although weighted models gave similar results. Fifth, cluster locations were moved slightly, and the travel-time map is from 2019 and measures travel to the nearest facility of any type. Both are likely to underestimate links with travel time. Sixth, mothers reported place of birth and caesarean themselves, and private facilities range from small clinics to large hospitals. Seventh, the nearby private share is an average over space. Placebo averages reduced clustering far less, but unmeasured local factors linked to private supply could still explain part of the effect. Eighth, clusters are small (median two facility births), so the maps show only broad regional patterns, and map probabilities are conditional on the model. Ninth, the surveys are cross-sectional and cover only each woman’s most recent live birth, not all births. First live births can still follow a caesarean after a pregnancy loss. Wealth, education and residence were measured after the birth, and women’s response was lower in 2012-13 (86.9%) than later.

### Implications

Facility birth and access to emergency obstetric care should keep expanding [35]. The benefits of more facility births may be partly lost if most facility births become surgical. Our results suggest the private sector is where reviews of caesarean use are most needed. High rates alone do not show how many caesareans were unnecessary; clinical audit is needed for that. Most private maternity care in Bangladesh is paid for by families themselves, and caesarean birth is linked to borrowing money and selling assets to pay for it [8]. Several measures deserve testing. These include licensing standards for private maternity units and required reporting of caesarean rates by Robson group, the ten-group classification of births by obstetric characteristics [5]. They also include payment arrangements that change provider incentives, clear prices, and stronger public emergency obstetric care [36]. Private-sector practice can change. In one Brazilian private hospital, women under a quality improvement model had a caesarean rate of 31.7%, against 88.6% under the standard model [37]. Severe maternal and neonatal outcomes did not clearly increase under that model. The central and western belt, where over 80% of facility births are caesarean, is a natural place to start. Public facilities offer a useful comparison within the same health system.

## Conclusions

In accounting terms, more births in facilities, especially private ones, made up most of the rise in caesareans in Bangladesh between 2012-13 and 2025. The net contribution of changing rates within sectors was small, but it combined a rising private rate with a falling public rate. The private caesarean rate rose beyond what measured case mix predicted, but among first births the excess was only about 5 to 6 points. Private facilities delivered most births by caesarean, for poor and rich women alike and at any modelled distance from a facility. If sector mix and sector rates stayed at 2025 levels, more facility births would keep raising the national caesarean rate. Expanding safe facility birth should go with monitoring of caesarean indications and quality of care.

## Declarations

### Funding statement

This research received no specific grant from any funding agency in the public, commercial or not-for-profit sectors.

### Competing interests

The authors have declared that no competing interests exist.

### Author contributions

FSH developed the study methodology, drafted the original manuscript, interpreted the findings, and critically reviewed and edited the manuscript. SMAUH conceptualized and designed the study, developed the analytical framework, and performed the formal analysis. SMAUH prepared the figures, validated the analytical results, coordinated the project, supervised the study, and managed the journal submission. SMAUH also contributed to methodological development and critically reviewed and edited the manuscript. MKI, MS, TN, FZ, USJ, and KN critically reviewed the findings and their interpretation and contributed to manuscript revision. All authors read and approved the final manuscript for submission.

## Data Availability

The MICS microdata and the 2025 cluster coordinates and covariates are available from the UNICEF MICS programme (https://mics.unicef.org) to registered users. Access is subject to the programme's terms of use.

## Acknowledgements

We thank the UNICEF MICS programme and the Bangladesh Bureau of Statistics for the survey data.

## Supporting information

**S1 Table. Validation against the published MICS 2012-13, 2019 and 2025 reports, place-of-birth harmonisation and missing data.**

**S2 Table. Within-sector prevalence ratios by round, all births and first births, private against public prevalence ratios, and E-values.**

**S3 Table. Decomposition by period: two-step and one-step versions, all births and first births, and illustrative scenarios.**

**S4 Table. Private-sector caesarean rate standardised for measured case mix, all births and first births, with paired differences.**

**S5 Table. Decomposition of the 2012-13 to 2025 change by wealth quintile and residence.**

**S6 Table. Caesarean rate within public and private facilities by wealth quintile and round.**

**S7 Table. Outcomes along the care pathway by band of modelled travel time to the nearest health facility, 2025: weighted percentages, adjusted prevalence ratios and sensitivity checks.**

**S8 Table. Spatial clustering of caesarean among facility births: survey-weighted models, placebo tests and sensitivity analyses, 2025.**

**S9 Table. Probability classes by division at thresholds 0.8 and 0.2, and 0.95 and 0.05, 2025.**

---

**S1 Fig.**
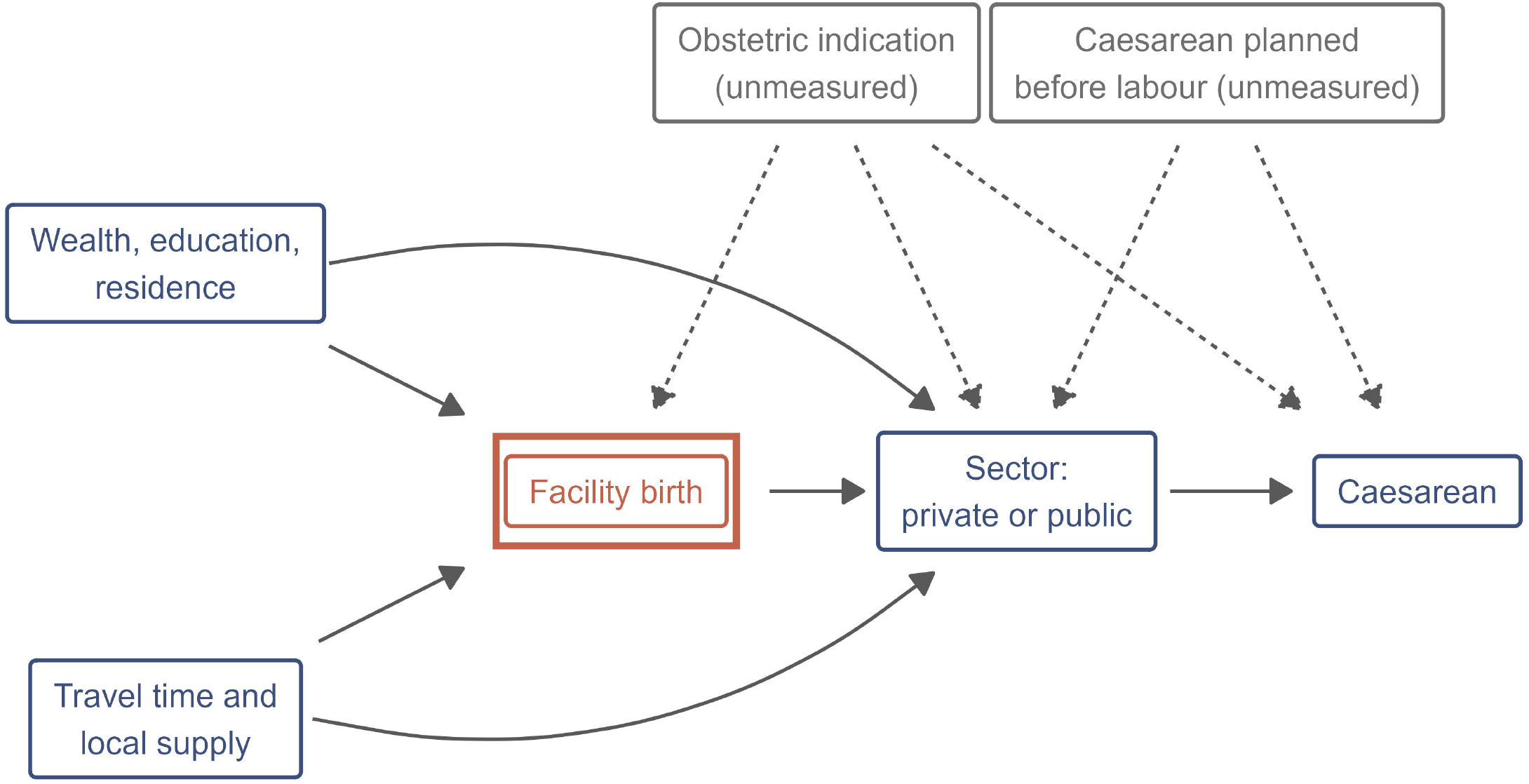
Directed acyclic graph of selection into facility and sector of birth.

**S2 Fig.**
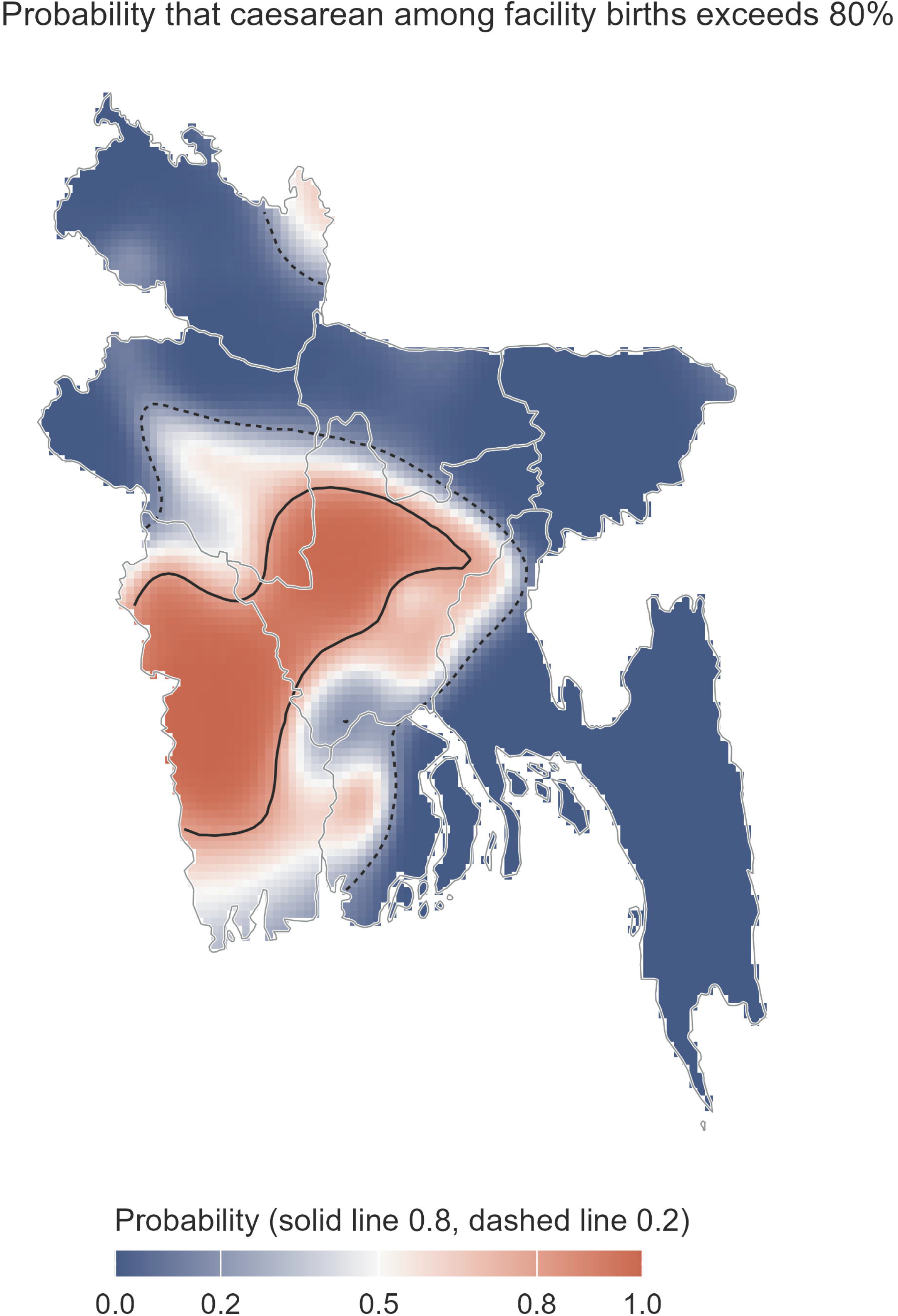
Probability that the caesarean rate among facility births exceeded 80%, 2025.

**S3 Fig.**
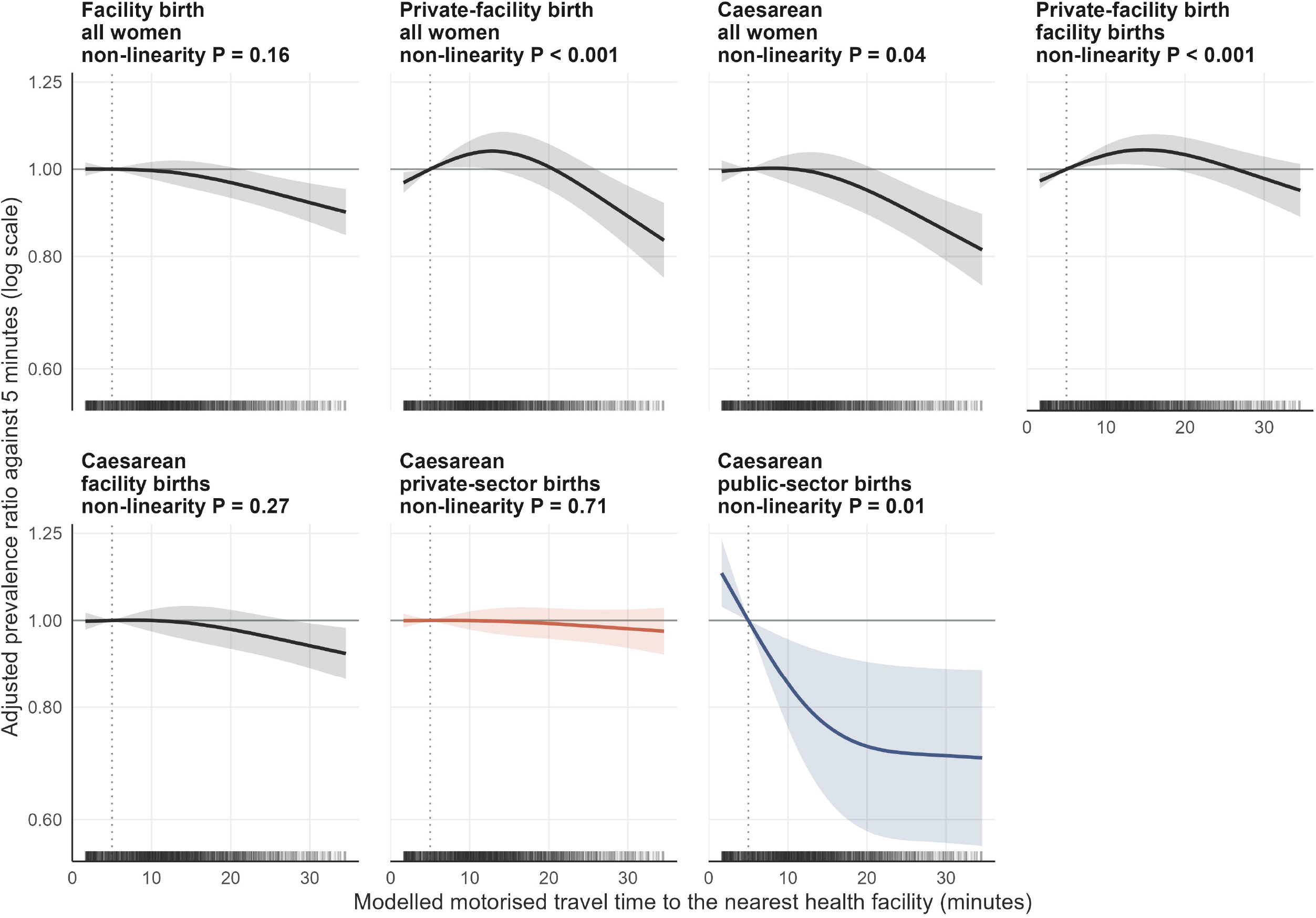
Adjusted prevalence ratio by modelled travel time to the nearest health facility, restricted cubic splines, 2025.

**S1 Checklist. STROBE checklist for cross-sectional studies.**

## Notes

### Competing Interest Statement

The authors have declared no competing interest.

### Author Declarations

This study used anonymised survey data from the UNICEF MICS programme. The 2025 survey protocol was approved by the icddr,b Ethical Review Committee (PR-24113). The 2019 protocol was approved by a government technical committee led by the Bangladesh Bureau of Statistics. The 2012-13 report does not describe its ethical review. The 2019 and 2025 surveys obtained verbal informed consent. We used cluster locations under the MICS data-use terms, only at cluster level, and made no attempt to identify households. No further ethical approval was needed for this secondary analysis.

