## Supplementary material for "Caesarean section in Bangladesh, 2012-13 to 2025: facility birth, the private sector and small-area geography": Supporting_information_v14.docx

### S1 Table. Validation against the published MICS reports, place-of-birth harmonisation and missing data

**A. Validation of weighted estimates against the published reports**

| Indicator | Round | This study (%) | Published (%) | Source |
| --- | --- | --- | --- | --- |
| Place of delivery: public facility | 2012-13 | 13.1 | 13.1 | MICS 2012-13 Table RH.11 |
| Place of delivery: private facility | 2012-13 | 17.9 | 17.9 | MICS 2012-13 Table RH.11 |
| Place of delivery: home | 2012-13 | 67.2 | 67.2 | MICS 2012-13 Table RH.11 |
| Place of delivery: other or missing | 2012-13 | 1.8 | 1.8 | MICS 2012-13 Table RH.11 |
| Delivered in a health facility | 2012-13 | 31.0 | 31.0 | MICS 2012-13 Table RH.11 |
| Caesarean section, report denominator | 2012-13 | 19.1 | 19.1 | MICS 2012-13 Table RH.10 |
| Caesarean section, analytic sample | 2012-13 | 19.5 |  | This study |
| Place of delivery: public facility | 2019 | 15.9 | 15.9 | MICS 2019 Table TM.6.1 |
| Place of delivery: private or NGO facility | 2019 | 37.5 | 37.5 | MICS 2019 Table TM.6.1 |
| Place of delivery: home | 2019 | 46.4 | 46.4 | MICS 2019 Table TM.6.1 |
| Delivered in a health facility | 2019 | 53.4 | 53.4 | MICS 2019 Table TM.6.1 |
| Caesarean section, report denominator | 2019 | 36.0 | 36.0 | MICS 2019 Table TM.6.2 |
| Caesarean section, analytic sample | 2019 | 36.1 |  | This study |
| Place of delivery: public facility | 2025 | 16.9 | 16.9 | MICS 2025 Table TM.6.1 |
| Place of delivery: private facility | 2025 | 52.8 | 52.8 | MICS 2025 Table TM.6.1 |
| Place of delivery: NGO facility | 2025 | 1.3 | 1.3 | MICS 2025 Table TM.6.1 |
| Place of delivery: unknown sector | 2025 | 0.1 | 0.1 | MICS 2025 Table TM.6.1 |
| Place of delivery: home | 2025 | 28.6 | 28.6 | MICS 2025 Table TM.6.1 |
| Place of delivery: other | 2025 | 0.2 | 0.2 | MICS 2025 Table TM.6.1 |
| Delivered in a health facility | 2025 | 71.2 | 71.0 | MICS 2025 Table TM.6.1 |
| Caesarean section, report denominator | 2025 | 51.8 | 51.8 | MICS 2025 Table TM.6.2 |
| Caesarean section, analytic sample | 2025 | 52.0 |  | This study |
| Caesarean decided before labour, all births | 2025 | 31.7 | 31.6 | MICS 2025 Table TM.6.2 |

*Place of delivery uses all women with place of delivery recorded (7,866, 9,285 and 9,542). “Report denominator” counts women with an unknown caesarean answer as not caesarean, as the reports do. “Analytic sample” excludes them, together with births at “other” places. The 2019 report places NGO facilities within the private sector. Private facilities include private maternity homes, as in the reports.*

**B. Harmonisation of place-of-birth categories (unweighted number of women)**

| Original survey category | Analysis class | 2012-13 | 2019 | 2025 |
| --- | --- | --- | --- | --- |
| Community clinic (CC) | Public | 0 | 101 | 107 |
| Government clinic / health centre | Public | 124 | 0 | 0 |
| Government clinic / health centre (FWC/USC/RD) | Public | 0 | 159 | 0 |
| Government clinic / health centre (UH&FWC/USC/RD) | Public | 0 | 0 | 230 |
| Government health post | Public | 4 | 0 | 0 |
| Government hospital | Public | 815 | 1,188 | 1,361 |
| Other public | Public | 10 | 5 | 15 |
| Other private | Private | 0 | 0 | 1 |
| Other private medical | Private | 25 | 0 | 0 |
| Private clinic | Private | 857 | 2,521 | 3,090 |
| Private hospital | Private | 344 | 690 | 1,719 |
| Private maternity home | Private | 18 | 46 | 42 |
| DK public, private, or NGO | NGO or unknown sector | 0 | 0 | 13 |
| NGO clinic | NGO or unknown sector | 0 | 0 | 59 |
| NGO clinic/hospital | NGO or unknown sector | 0 | 105 | 0 |
| NGO hospital | NGO or unknown sector | 0 | 0 | 34 |
| Other NGO | NGO or unknown sector | 0 | 0 | 4 |
| Other home | Home | 480 | 730 | 340 |
| Respondent’s home | Home | 5,080 | 3,726 | 2,504 |
| Missing | Excluded (other or missing) | 71 | 0 | 0 |
| No response | Excluded (other or missing) | 0 | 1 | 0 |
| Other | Excluded (other or missing) | 38 | 13 | 23 |

*Category labels are as recorded in each round’s data file. The 2012-13 survey had no separate NGO category. Births with a recorded caesarean occur only in facility categories.*

**C. Missing values in the analytic sample**

| Round | Women | Age group | Parity | Education | Wealth | Residence | Division | Four or more antenatal visits | Complete on all covariates |
| --- | --- | --- | --- | --- | --- | --- | --- | --- | --- |
| 2012-13 | 7,750 | 0 | 0 | 0 | 0 | 0 | 0 | 37 | 7,713 |
| 2019 | 9,270 | 0 | 0 | 0 | 0 | 0 | 0 | 5 | 9,265 |
| 2025 | 9,519 | 0 | 0 | 4 | 0 | 0 | 0 | 21 | 9,494 |

*Antenatal care is missing when the number of visits was “don’t know” or the any-care question was unanswered; women with no antenatal care count as fewer than four visits. Models use complete cases. Modelled travel time was available for every 2025 cluster.*

### S2 Table. Within-sector prevalence ratios by round, all births and first births, private against public prevalence ratios, and E-values

| Sector | Model | Contrast | PR (95% CI) | n |
| --- | --- | --- | --- | --- |
| Public | Crude | 2019 vs 2012-13 | 0.91 (0.79 to 1.05) | 4,117 |
| Public | Crude | 2025 vs 2012-13 | 0.87 (0.75 to 1.00) | 4,117 |
| Public | Adjusted | 2019 vs 2012-13 | 0.94 (0.82 to 1.07) | 4,111 |
| Public | Adjusted | 2025 vs 2012-13 | 0.94 (0.82 to 1.08) | 4,111 |
| Private | Crude | 2019 vs 2012-13 | 1.05 (1.00 to 1.10) | 9,347 |
| Private | Crude | 2025 vs 2012-13 | 1.09 (1.04 to 1.14) | 9,347 |
| Private | Adjusted | 2019 vs 2012-13 | 1.06 (1.01 to 1.11) | 9,324 |
| Private | Adjusted | 2025 vs 2012-13 | 1.10 (1.06 to 1.15) | 9,324 |

| Round | Private vs public, adjusted PR (95% CI) | n |
| --- | --- | --- |
| 2012-13 | 1.87 (1.65 to 2.12) | 2,180 |
| 2019 | 2.26 (2.08 to 2.46) | 4,706 |
| 2025 | 2.46 (2.27 to 2.67) | 6,549 |

**First births only (parity 1), adjusted**

| Sector | Contrast | PR (95% CI) | n |
| --- | --- | --- | --- |
| Public | 2019 vs 2012-13 | 0.87 (0.71 to 1.06) | 1,716 |
| Public | 2025 vs 2012-13 | 0.95 (0.77 to 1.15) | 1,716 |
| Private | 2019 vs 2012-13 | 1.04 (0.99 to 1.11) | 4,038 |
| Private | 2025 vs 2012-13 | 1.06 (1.00 to 1.12) | 4,038 |

**First births only: weighted percentages by round (95% CI)**

| Measure | 2012-13 | 2019 | 2025 |
| --- | --- | --- | --- |
| Caesarean, public-sector first births | 42.1 (35.1 to 49.1) | 32.5 (28.2 to 36.8) | 35.5 (31.3 to 39.7) |
| Caesarean, private-sector first births | 81.7 (77.2 to 86.2) | 85.0 (82.8 to 87.1) | 86.1 (84.4 to 87.9) |
| Facility birth, first births | 42.1 (39.3 to 44.8) | 64.8 (62.8 to 66.7) | 79.5 (77.9 to 81.1) |
| Caesarean, all first births | 28.0 (25.4 to 30.7) | 44.5 (42.5 to 46.6) | 58.6 (56.7 to 60.6) |

**E-values for the 2025 against 2012-13 contrasts (adjusted)**

| Sector and births | PR (95% CI) | E-value, estimate | E-value, confidence limit closest to 1 |
| --- | --- | --- | --- |
| Public all births | 0.94 (0.82 to 1.08) | 1.32 | 1.00 |
| Private all births | 1.10 (1.06 to 1.15) | 1.44 | 1.30 |
| Public first births | 0.95 (0.77 to 1.15) | 1.31 | 1.00 |
| Private first births | 1.06 (1.00 to 1.12) | 1.32 | 1.07 |

*E-value: the minimum risk ratio that an unmeasured factor would need with both survey round and caesarean, beyond the measured covariates, to explain away the estimate (VanderWeele and Ding, 2017). An E-value of 1.00 for the confidence limit means the interval already includes 1. Post hoc analysis.*

*Survey-weighted Poisson models with design-based (Taylor linearisation) variance; survey round entered as a categorical term. Adjusted models include age group, parity, wealth quintile, residence, education, division and four or more antenatal visits. First-birth models omit parity. A first birth cannot follow a previous caesarean, so this comparison removes repeat caesarean.*

### S3 Table. Decomposition by period: two-step and one-step versions, all births and first births, and illustrative scenarios

**Two-step decomposition (percentage points, 95% CI)**

| Component | 2012-13 to 2019 | 2019 to 2025 | 2012-13 to 2025 |
| --- | --- | --- | --- |
| Total change | 16.6 (14.6 to 18.4) | 15.9 (14.2 to 17.7) | 32.5 (30.6 to 34.4) |
| More births in facilities | 14.2 (12.8 to 15.7); 86.0% | 12.5 (11.3 to 13.8); 78.7% | 26.9 (25.4 to 28.4); 82.8% |
| Shift from public to private facilities | 1.8 (1.1 to 2.5); 10.9% | 2.1 (1.4 to 2.8); 13.1% | 4.1 (3.2 to 4.8); 12.6% |
| Change in unadjusted rates within sectors | 0.5 (-0.9 to 1.8); 3.1% | 1.3 (0.2 to 2.5); 8.2% | 1.5 (0.0 to 3.0); 4.6% |

**One-step decomposition over four places of birth**

| Period | Total change | Composition (place of birth) | Within-place rates |
| --- | --- | --- | --- |
| 2012-13 to 2019 | 16.6 (14.7 to 18.5) | 16.0 (14.6 to 17.6); 96.7% (89.9 to 104.5) | 0.6 (-0.7 to 1.8); 3.3% (-4.5 to 10.1) |
| 2019 to 2025 | 15.9 (14.0 to 17.7) | 14.6 (13.1 to 16.0); 91.7% (86.3 to 98.6) | 1.3 (0.2 to 2.3); 8.3% (1.4 to 13.7) |
| 2012-13 to 2025 | 32.5 (30.5 to 34.3) | 30.7 (29.2 to 32.3); 94.7% (90.7 to 98.9) | 1.7 (0.4 to 3.1); 5.3% (1.1 to 9.3) |
| First births only, 2012-13 to 2025 | 30.6 (27.4 to 33.8) | 29.8 (27.2 to 32.2); 97.4% | 0.8 (-1.7 to 3.4); 2.6% |

*Percentage points (95% CI) and share of the total change. Shares are accounting contributions under each decomposition, not causal fractions. One-step places: home, public, private, NGO or unknown. The two-step version splits composition into facility birth and sector mix; that split depends on the order of the steps. Intervals from 1,000 Rao-Wu rescaling bootstrap replicates of clusters within strata; every component is recomputed in each replicate.*

**Within-sector term split by sector (percentage points, 95% CI)**

| Sector | 2012-13 to 2019 | 2019 to 2025 | 2012-13 to 2025 |
| --- | --- | --- | --- |
| Public | -0.5 (-1.3 to 0.4) | -0.3 (-0.8 to 0.3) | -0.9 (-1.7 to 0.0) |
| Private | 1.1 (0.1 to 2.0) | 1.4 (0.5 to 2.2) | 2.4 (1.3 to 3.6) |
| NGO or unknown | 0.0 (0.0 to 0.0) | 0.2 (0.0 to 0.4) | 0.0 (0.0 to 0.0) |
| Total within-sector term | 0.5 (-0.7 to 1.8) | 1.3 (0.2 to 2.4) | 1.5 (0.1 to 2.9) |

*Each cell is the average facility share times the average sector share times the change in that sector’s caesarean rate. Separate bootstrap run (1,000 replicates), so the total differs slightly in its interval from the two-step table above. The NGO category did not exist in 2012-13, so its 2012-13 rate is set to its later rate.*

**Illustrative scenarios for the national caesarean rate, 2025 inputs (percent, 95% CI)**

| Scenario | Rate |
| --- | --- |
| Observed 2025 | 52.0 (50.8 to 53.2) |
| Facility birth 90%, 2025 sector mix and sector rates | 65.5 (64.4 to 66.8) |
| Facility birth 90%, extra facility births in public facilities at the 2025 public rate | 58.2 (56.9 to 59.5) |

*Scenarios hold sector rates fixed. Intervals come from 1,000 bootstrap replicates, so the observed interval differs slightly from the linearised interval in Table 2.*

### S4 Table. Private-sector caesarean rate standardised for measured case mix, all births and first births, with paired differences

**A. All private births (upper bound, because previous caesarean is not recorded)**

| Model fitted to round | Case mix 2012-13 | Case mix 2019 | Case mix 2025 |
| --- | --- | --- | --- |
| 2012-13 | **79.0 (75.5 to 81.9)** | 77.6 (74.0 to 80.9) | 76.8 (72.9 to 80.5) |
| 2019 | 83.6 (81.8 to 85.4) | **83.0 (81.4 to 84.5)** | 82.5 (80.9 to 84.2) |
| 2025 | 86.9 (85.3 to 88.5) | 86.3 (85.1 to 87.6) | **86.1 (84.9 to 87.3)** |

| Comparison | Difference (percentage points, 95% CI) |
| --- | --- |
| Observed 2025 minus 2012-13 model applied to the 2025 case mix | 9.3 (5.4 to 13.4) |
| 2025 model applied to the 2012-13 case mix minus observed 2012-13 | 7.9 (4.3 to 11.7) |

**B. Private first births only (main estimate; no previous caesarean possible)**

| Model fitted to round | Case mix 2012-13 | Case mix 2019 | Case mix 2025 |
| --- | --- | --- | --- |
| 2012-13 | **81.2 (76.0 to 85.3)** | 81.6 (76.8 to 85.4) | 81.3 (76.1 to 85.5) |
| 2019 | 85.1 (82.0 to 87.6) | **84.9 (82.6 to 87.0)** | 84.7 (82.3 to 86.8) |
| 2025 | 87.3 (84.9 to 89.5) | 86.7 (84.8 to 88.4) | **86.1 (84.3 to 87.8)** |

| Comparison | Difference (percentage points, 95% CI) |
| --- | --- |
| Observed 2025 minus 2012-13 model applied to the 2025 case mix | 4.8 (0.3 to 10.2) |
| 2025 model applied to the 2012-13 case mix minus observed 2012-13 | 6.1 (1.2 to 11.6) |

**C. Private-sector case mix (weighted percent)**

| Round | First births | Poorest two quintiles | Rural | Age 30 or older | Fewer than four antenatal visits |
| --- | --- | --- | --- | --- | --- |
| 2012-13 | 52.4 | 15.1 | 64.0 | 21.9 | 46.2 |
| 2019 | 42.7 | 21.1 | 72.8 | 24.4 | 45.5 |
| 2025 | 43.0 | 28.6 | 70.6 | 24.0 | 45.0 |

*A separate Poisson model is fitted to each round’s private births, so every coefficient may differ between rounds. Each model is then applied to the private births of every round (marginal standardisation). Bold cells are observed rates in the standardisation sample, which excludes women with unknown antenatal care. Models include age group, parity (not in panel B), wealth quintile, residence, education, division and four or more antenatal visits. Models are refitted in each of 1,000 bootstrap replicates, and differences are computed within each replicate, so intervals include model-fitting uncertainty. Prediction was impossible for at most 2 women per comparison because a covariate level was absent in the fitting round. Clinical factors such as presentation, multiple pregnancy, gestational age and complications are not recorded, so differences describe caesarean use not explained by measured case mix. Omitting antenatal care gave 2025 gaps of 9.5 points (all births) and 4.3 points (first births). Wealth quintiles rank households within each round.*

*Unweighted caesareans among private first births with known antenatal care: 2012-13, 504 of 607; 2019, 1,182 of 1,385; 2025, 1,773 of 2,046.*

### S5 Table. Decomposition of the 2012-13 to 2025 change by wealth quintile and residence

| Group | Rate 2012-13 (%) | Rate 2025 (%) | Total change (points) | More births in facilities | Shift from public to private | Change in rates within sectors |
| --- | --- | --- | --- | --- | --- | --- |
| Poorest quintile | 5.5 (4.5 to 6.7) | 34.3 (32.0 to 36.9) | 28.8 (26.2 to 31.6) | 21.3 (19.2 to 23.6); 73.8% of change | 5.9 (4.3 to 7.4); 20.3% of change | 1.7 (-0.6 to 3.9); 5.8% of change |
| Quintile 2 | 8.8 (7.3 to 10.5) | 45.1 (42.4 to 48.0) | 36.2 (33.0 to 39.5) | 27.9 (25.0 to 31.1); 76.9% of change | 4.9 (3.5 to 6.6); 13.6% of change | 3.4 (0.0 to 6.6); 9.5% of change |
| Quintile 3 | 13.0 (10.9 to 15.0) | 52.2 (49.6 to 54.5) | 39.2 (35.9 to 42.7) | 30.2 (27.4 to 33.0); 76.9% of change | 6.5 (4.5 to 8.8); 16.6% of change | 2.5 (-0.6 to 5.8); 6.4% of change |
| Quintile 4 | 25.8 (22.8 to 28.7) | 57.4 (54.9 to 60.2) | 31.6 (27.6 to 35.7) | 25.5 (22.5 to 28.8); 80.7% of change | 4.7 (2.8 to 6.4); 14.8% of change | 1.4 (-1.8 to 4.6); 4.4% of change |
| Richest quintile | 47.3 (42.6 to 51.6) | 68.3 (65.5 to 71.0) | 21.0 (16.0 to 26.1) | 16.2 (12.9 to 19.6); 77.3% of change | 2.8 (1.2 to 4.5); 13.2% of change | 2.0 (-2.0 to 5.9); 9.5% of change |
| Urban | 33.9 (29.1 to 39.0) | 55.6 (53.1 to 58.1) | 21.8 (16.1 to 27.1) | 17.3 (13.7 to 21.2); 79.3% of change | 2.9 (1.2 to 4.7); 13.4% of change | 1.6 (-2.5 to 5.3); 7.3% of change |
| Rural | 15.7 (14.4 to 16.9) | 50.5 (49.1 to 51.9) | 34.8 (32.9 to 36.9) | 28.7 (27.2 to 30.2); 82.3% of change | 4.7 (3.8 to 5.5); 13.4% of change | 1.5 (0.1 to 2.9); 4.3% of change |

*Two-step decomposition, percentage points (95% CI) from 1,000 bootstrap replicates. Wealth quintiles rank households within each round.*

### S6 Table. Caesarean rate within public and private facilities, and private caesarean among all recent mothers, by wealth quintile and round

| Sector | Wealth quintile | 2012-13 | 2019 | 2025 |
| --- | --- | --- | --- | --- |
| Private | Poorest | 74.4 (64.3 to 84.6), n 99 | 77.2 (71.5 to 82.8), n 301 | 86.3 (83.4 to 89.2), n 703 |
| Private | Second | 73.1 (62.6 to 83.5), n 134 | 83.4 (79.7 to 87.1), n 488 | 85.7 (82.9 to 88.5), n 879 |
| Private | Middle | 80.0 (71.6 to 88.4), n 186 | 83.6 (80.4 to 86.7), n 679 | 86.4 (83.7 to 89.1), n 1,028 |
| Private | Fourth | 77.1 (71.0 to 83.2), n 309 | 83.6 (80.5 to 86.8), n 821 | 87.0 (84.5 to 89.4), n 1,087 |
| Private | Richest | 81.4 (76.7 to 86.1), n 510 | 83.5 (80.9 to 86.1), n 968 | 85.3 (82.8 to 87.7), n 1,155 |
| Public | Poorest | 23.4 (16.2 to 30.5), n 175 | 22.3 (16.5 to 28.1), n 267 | 21.2 (16.3 to 26.1), n 366 |
| Public | Second | 30.3 (22.1 to 38.4), n 164 | 31.1 (25.4 to 36.9), n 301 | 32.8 (27.0 to 38.6), n 378 |
| Public | Middle | 29.3 (20.9 to 37.7), n 176 | 33.6 (27.3 to 39.9), n 280 | 32.7 (26.9 to 38.5), n 365 |
| Public | Fourth | 45.0 (37.1 to 52.9), n 205 | 34.5 (29.0 to 40.0), n 341 | 32.1 (26.3 to 37.9), n 345 |
| Public | Richest | 50.9 (39.9 to 61.8), n 232 | 49.5 (42.5 to 56.4), n 263 | 49.7 (42.6 to 56.7), n 259 |

*Weighted percent (95% CI) and unweighted number of births in the sector.*

*Test of wealth differences in 2025 (survey-weighted Poisson models, unadjusted): private facilities, richest against poorest PR 0.99 (0.95 to 1.03), Wald test of any wealth difference P = 0.90; public facilities, PR 2.34 (1.79 to 3.08), P < 0.001. Wealth by sector interaction P < 0.001. Adjusted for age group, parity, education, residence and division: private facilities, PR 1.03 (0.98 to 1.08), P = 0.57; public facilities, PR 2.18 (1.61 to 2.95), P < 0.001.*

**Among all recent mothers (not only facility births): weighted percent (95% CI) by wealth quintile**

| Outcome | Round | Poorest | Second | Middle | Fourth | Richest |
| --- | --- | --- | --- | --- | --- | --- |
| Birth in a private facility | 2012-13 | 4.8 (3.8 to 5.9) | 8.1 (6.5 to 9.6) | 11.7 (9.8 to 13.7) | 23.3 (20.4 to 26.2) | 45.5 (41.1 to 50.0) |
| Birth in a private facility | 2019 | 13.5 (11.8 to 15.2) | 25.2 (22.9 to 27.4) | 36.8 (34.3 to 39.3) | 45.2 (42.5 to 48.0) | 59.5 (56.5 to 62.4) |
| Birth in a private facility | 2025 | 35.4 (33.0 to 37.9) | 45.0 (42.3 to 47.8) | 53.5 (50.9 to 56.1) | 58.4 (55.8 to 61.1) | 69.7 (67.2 to 72.2) |
| Caesarean in a private facility | 2012-13 | 3.6 (2.7 to 4.5) | 5.9 (4.6 to 7.2) | 9.4 (7.7 to 11.1) | 17.9 (15.3 to 20.5) | 37.1 (32.8 to 41.3) |
| Caesarean in a private facility | 2019 | 10.4 (9.0 to 11.9) | 21.0 (18.9 to 23.1) | 30.7 (28.4 to 33.1) | 37.8 (35.1 to 40.5) | 49.6 (46.6 to 52.7) |
| Caesarean in a private facility | 2025 | 30.6 (28.2 to 32.9) | 38.6 (36.1 to 41.1) | 46.2 (43.6 to 48.9) | 50.8 (48.2 to 53.5) | 59.4 (56.6 to 62.2) |
| Caesarean, any place | 2012-13 | 5.5 (4.4 to 6.7) | 8.8 (7.3 to 10.4) | 13.0 (11.0 to 15.0) | 25.8 (22.8 to 28.8) | 47.3 (42.9 to 51.7) |
| Caesarean, any place | 2019 | 13.2 (11.6 to 14.8) | 26.1 (23.8 to 28.3) | 36.5 (34.1 to 39.0) | 44.6 (41.8 to 47.3) | 59.6 (56.7 to 62.5) |
| Caesarean, any place | 2025 | 34.3 (31.9 to 36.8) | 45.1 (42.4 to 47.7) | 52.2 (49.6 to 54.8) | 57.4 (54.8 to 59.9) | 68.3 (65.5 to 71.0) |

*In 2025, richest against poorest, private caesarean among all recent mothers: PR 1.94 (1.78 to 2.13), unadjusted survey-weighted Poisson model. This separates reaching a private facility from caesarean once there.*

### S7 Table. Outcomes along the care pathway by band of modelled travel time to the nearest health facility of any type, 2025

| Outcome | Population | Band (minutes) | Weighted % (95% CI) | Adjusted PR (95% CI) | Also adjusted for division |
| --- | --- | --- | --- | --- | --- |
| Birth in a facility | All women | <5 | 78.6 (76.1 to 81.1) | 1 (reference) | 1 (reference) |
| Birth in a facility | All women | 5 to <10 | 73.5 (70.9 to 76.1) | 1.03 (0.98 to 1.09) | 1.03 (0.98 to 1.09) |
| Birth in a facility | All women | 10 to <20 | 71.4 (69.5 to 73.3) | 1.04 (0.99 to 1.09) | 1.02 (0.97 to 1.08) |
| Birth in a facility | All women | 20 to <30 | 60.4 (56.7 to 64.2) | 0.91 (0.85 to 0.98) | 0.89 (0.83 to 0.95) |
| Birth in a facility | All women | 30+ | 57.4 (51.4 to 63.4) | 0.89 (0.81 to 0.98) | 0.89 (0.80 to 0.98) |
| Birth in a private facility | All women | <5 | 58.3 (55.3 to 61.3) | 1 (reference) | 1 (reference) |
| Birth in a private facility | All women | 5 to <10 | 54.4 (51.6 to 57.1) | 1.04 (0.96 to 1.13) | 1.08 (1.00 to 1.17) |
| Birth in a private facility | All women | 10 to <20 | 54.3 (52.2 to 56.5) | 1.09 (1.01 to 1.18) | 1.13 (1.04 to 1.22) |
| Birth in a private facility | All women | 20 to <30 | 43.6 (40.3 to 46.8) | 0.92 (0.83 to 1.02) | 0.94 (0.85 to 1.04) |
| Birth in a private facility | All women | 30+ | 37.9 (32.3 to 43.5) | 0.83 (0.72 to 0.97) | 0.89 (0.77 to 1.02) |
| Caesarean | All women | <5 | 60.8 (57.8 to 63.7) | 1 (reference) | 1 (reference) |
| Caesarean | All women | 5 to <10 | 52.1 (49.4 to 54.8) | 0.94 (0.87 to 1.03) | 1.00 (0.92 to 1.08) |
| Caesarean | All women | 10 to <20 | 51.9 (49.8 to 54.1) | 0.98 (0.90 to 1.07) | 1.03 (0.95 to 1.11) |
| Caesarean | All women | 20 to <30 | 42.7 (39.3 to 46.2) | 0.85 (0.76 to 0.95) | 0.87 (0.78 to 0.96) |
| Caesarean | All women | 30+ | 37.2 (31.7 to 42.7) | 0.77 (0.67 to 0.89) | 0.83 (0.72 to 0.96) |
| Birth in a private facility | Facility births | <5 | 74.2 (71.6 to 76.8) | 1 (reference) | 1 (reference) |
| Birth in a private facility | Facility births | 5 to <10 | 73.9 (71.3 to 76.6) | 1.01 (0.95 to 1.08) | 1.05 (0.99 to 1.12) |
| Birth in a private facility | Facility births | 10 to <20 | 76.1 (74.0 to 78.1) | 1.06 (0.99 to 1.12) | 1.10 (1.04 to 1.17) |
| Birth in a private facility | Facility births | 20 to <30 | 72.1 (68.5 to 75.6) | 1.01 (0.94 to 1.09) | 1.05 (0.98 to 1.13) |
| Birth in a private facility | Facility births | 30+ | 66.0 (58.9 to 73.0) | 0.93 (0.83 to 1.04) | 0.99 (0.88 to 1.11) |
| Caesarean | Facility births | <5 | 77.3 (74.5 to 80.2) | 1 (reference) | 1 (reference) |
| Caesarean | Facility births | 5 to <10 | 70.9 (68.1 to 73.8) | 0.92 (0.85 to 1.00) | 0.98 (0.91 to 1.05) |
| Caesarean | Facility births | 10 to <20 | 72.7 (70.5 to 74.9) | 0.95 (0.88 to 1.03) | 1.01 (0.94 to 1.08) |
| Caesarean | Facility births | 20 to <30 | 70.7 (67.0 to 74.5) | 0.93 (0.86 to 1.02) | 0.97 (0.89 to 1.05) |
| Caesarean | Facility births | 30+ | 64.8 (57.9 to 71.7) | 0.86 (0.76 to 0.97) | 0.93 (0.82 to 1.04) |
| Caesarean | Private-sector births | <5 | 87.6 (85.1 to 90.1) | 1 (reference) | 1 (reference) |
| Caesarean | Private-sector births | 5 to <10 | 84.5 (81.8 to 87.2) | 0.95 (0.90 to 1.01) | 0.98 (0.93 to 1.03) |
| Caesarean | Private-sector births | 10 to <20 | 86.3 (84.2 to 88.3) | 0.97 (0.92 to 1.02) | 0.99 (0.94 to 1.05) |
| Caesarean | Private-sector births | 20 to <30 | 86.5 (83.2 to 89.8) | 0.97 (0.91 to 1.03) | 0.98 (0.92 to 1.04) |
| Caesarean | Private-sector births | 30+ | 83.7 (76.5 to 90.9) | 0.93 (0.85 to 1.03) | 0.95 (0.86 to 1.05) |
| Caesarean | Public-sector births | <5 | 47.9 (41.5 to 54.2) | 1 (reference) | 1 (reference) |
| Caesarean | Public-sector births | 5 to <10 | 31.1 (25.7 to 36.5) | 0.68 (0.52 to 0.89) | 0.74 (0.57 to 0.97) |
| Caesarean | Public-sector births | 10 to <20 | 29.2 (24.8 to 33.6) | 0.63 (0.48 to 0.84) | 0.69 (0.52 to 0.91) |
| Caesarean | Public-sector births | 20 to <30 | 27.9 (20.9 to 34.9) | 0.63 (0.44 to 0.89) | 0.65 (0.46 to 0.92) |
| Caesarean | Public-sector births | 30+ | 25.6 (16.4 to 34.8) | 0.59 (0.39 to 0.89) | 0.71 (0.48 to 1.05) |

**Sensitivity checks (per 10 minutes, or per hour of walking time)**

| Outcome | Population | Check | Adjusted PR (95% CI) | n |
| --- | --- | --- | --- | --- |
| Birth in a facility | All women | Under 60 minutes | 0.95 (0.93 to 0.98) | 9,421 |
| Birth in a facility | All women | Walking time, per hour | 0.95 (0.93 to 0.98) | 9,515 |
| Caesarean | All women | Under 60 minutes | 0.94 (0.91 to 0.97) | 9,421 |
| Caesarean | All women | Walking time, per hour | 0.93 (0.90 to 0.96) | 9,515 |
| Caesarean | Facility births | Under 60 minutes | 0.98 (0.95 to 1.00) | 6,630 |
| Caesarean | Facility births | Walking time, per hour | 0.97 (0.95 to 1.00) | 6,673 |

*Travel time is modelled motorised travel time from the displaced cluster point to the nearest health facility of any type (2019 map), not to a facility offering caesarean or emergency obstetric care. Models adjust for age group, parity, wealth quintile, residence and education. Facility-birth models include 6,673 of 6,675 facility births; two lacked education data. The per-10-minute PRs in Table 3 summarise patterns that are not always linear; the bands show their shape.*

*Weighted median travel time was 11.0 minutes, and 6.0% of the 9,519 women lived 30 minutes or more away.*

**Unweighted events and denominators by travel-time band**

| Outcome | Population | <5 | 5 to <10 | 10 to <20 | 20 to <30 | 30+ |
| --- | --- | --- | --- | --- | --- | --- |
| Birth in a facility | All women | 1,292/1,628 | 1,487/2,029 | 2,664/3,749 | 832/1,374 | 400/739 |
| Birth in a private facility | All women | 951/1,628 | 1,076/2,029 | 1,995/3,749 | 590/1,374 | 240/739 |
| Caesarean | All women | 1,002/1,628 | 1,062/2,029 | 1,958/3,749 | 581/1,374 | 236/739 |
| Birth in a private facility | Facility births | 951/1,292 | 1,076/1,487 | 1,995/2,664 | 590/832 | 240/400 |
| Caesarean | Facility births | 1,002/1,292 | 1,062/1,487 | 1,958/2,664 | 581/832 | 236/400 |
| Caesarean | Private-sector births | 844/951 | 924/1,076 | 1,750/1,995 | 515/590 | 196/240 |
| Caesarean | Public-sector births | 136/299 | 128/394 | 195/640 | 60/230 | 35/150 |

### S8 Table. Spatial clustering of caesarean among facility births: survey-weighted models, placebo tests and sensitivity analyses, 2025

**A. Survey-weighted models**

| Cluster-level model | Moran’s I | Permutation P |
| --- | --- | --- |
| No covariates | 0.185 | 0.001 |
| Household factors | 0.195 | 0.001 |
| Plus travel time | 0.187 | 0.001 |
| Plus neighbouring delivery-sector composition (private share) | 0.090 | 0.001 |
| Plus division | 0.045 | 0.001 |
| Alternative: district private share, own cluster excluded | 0.083 | 0.001 |
| Alternative: own-cluster private share | 0.140 | 0.001 |

*Quasi-binomial models of the survey-weighted cluster rate with normalised weights; 2,720 clusters, eight nearest neighbours, 999 permutations. Unweighted results are in Table 4 of the main text.*

**B. Placebo tests, unweighted binomial models (Moran’s I, analytic)**

| Model | Moran’s I |
| --- | --- |
| Household factors and travel time | 0.183 |
| Plus neighbouring private share | 0.070 |
| Plus neighbours’ mean elevation | 0.176 |
| Plus neighbours’ mean rainfall, 2020 | 0.155 |
| Plus neighbours’ mean wealth quintile | 0.172 |
| Plus neighbours’ average of permuted private share, median of 200 permutations | 0.183 (range 0.176 to 0.184) |

*Placebo covariates are averaged over the same eight nearest neighbours. Elevation and rainfall come from the MICS 2025 cluster covariate file. The permuted share has the same averaging but no geographic structure, so it is a weaker placebo than elevation and rainfall.*

**C. Number of neighbours (neighbour weights and private share use the same number)**

| Neighbours | Moran’s I, household factors and travel time | Moran’s I, plus neighbouring private share | OR per 10-point higher share (95% CI) |
| --- | --- | --- | --- |
| 4 | 0.191 | 0.077 | 1.21 (1.17 to 1.24) |
| 8 | 0.183 | 0.070 | 1.30 (1.26 to 1.35) |
| 12 | 0.180 | 0.061 | 1.38 (1.32 to 1.43) |

**D. Basis dimension of the spatial smooth**

| Basis dimension | Effective df, unadjusted surface | Clusters with P > 0.8 above national rate | Clusters with P < 0.2 | Moran’s I, full model | Barishal, % of clusters above | Chattogram, % of clusters above | Dhaka, % of clusters above | Khulna, % of clusters above | Mymensingh, % of clusters above | Rajshahi, % of clusters above | Rangpur, % of clusters above | Sylhet, % of clusters above |
| --- | --- | --- | --- | --- | --- | --- | --- | --- | --- | --- | --- | --- |
| 40 | 26.3 | 1,497 | 709 | 0.015 | 60 | 16 | 97 | 100 | 25 | 53 | 15 | 0 |
| 80 | 37.1 | 1,486 | 674 | 0.009 | 65 | 16 | 96 | 100 | 12 | 55 | 12 | 0 |
| 120 | 40.5 | 1,475 | 678 | 0.008 | 65 | 15 | 96 | 100 | 10 | 56 | 11 | 0 |

**E. Leaving out one division at a time**

| Division left out | Clusters | Moran’s I, household factors and travel time | Moran’s I, plus neighbouring private share | OR per 10-point higher share (95% CI) |
| --- | --- | --- | --- | --- |
| Barishal | 2,485 | 0.185 | 0.056 | 1.32 (1.27 to 1.37) |
| Chattogram | 2,239 | 0.079 | 0.030 | 1.19 (1.13 to 1.24) |
| Dhaka | 2,100 | 0.186 | 0.078 | 1.30 (1.25 to 1.35) |
| Khulna | 2,302 | 0.186 | 0.081 | 1.28 (1.23 to 1.33) |
| Mymensingh | 2,564 | 0.195 | 0.076 | 1.32 (1.27 to 1.37) |
| Rajshahi | 2,385 | 0.188 | 0.076 | 1.31 (1.26 to 1.36) |
| Rangpur | 2,405 | 0.195 | 0.072 | 1.33 (1.28 to 1.38) |
| Sylhet | 2,560 | 0.196 | 0.075 | 1.32 (1.27 to 1.37) |

**F. Facility births per cluster**

| Facility births in cluster | 1 | 2 | 3 | 4 to 5 | 6 or more |
| --- | --- | --- | --- | --- | --- |
| Clusters | 801 | 769 | 602 | 463 | 85 |

*Analytic Moran’s I in panels C to E (no permutation). Clusters hold a median of 2 facility births, so smoothed values borrow strength from neighbouring clusters. Probabilities in panel D are conditional on the fitted model; they do not include survey design, coordinate displacement or model choice.*

**G. Distance to the eight nearest clusters (km)**

| Residence | Clusters | Median of mean distance | 10th to 90th centile | Median distance to farthest of the eight |
| --- | --- | --- | --- | --- |
| Rural | 1,979 | 7.3 | 4.6 to 11.3 | 10.4 |
| Urban | 741 | 5.8 | 1.9 to 10.8 | 8.8 |

**H. Distance-band neighbours (all clusters within the radius)**

| Radius (km) | Clusters | Clusters with no neighbour (dropped) | Median neighbours | Moran’s I, household factors and travel time | Moran’s I, plus neighbouring private share | OR per 10-point higher share (95% CI) |
| --- | --- | --- | --- | --- | --- | --- |
| 10 | 2,687 | 33 | 7 | 0.194 | 0.071 | 1.22 (1.19 to 1.26) |
| 15 | 2,716 | 4 | 16 | 0.179 | 0.058 | 1.35 (1.30 to 1.41) |
| 20 | 2,719 | 1 | 27 | 0.166 | 0.041 | 1.44 (1.38 to 1.50) |

*Analytic Moran’s I with row-standardised distance-band weights. Odds ratios exaggerate prevalence ratios for this common outcome and are shown only to compare specifications.*

*Pearson dispersion of the unweighted binomial model with the neighbouring private share was 1.07. Across the 5th to 95th centiles of the grid, odds ratios of the spatial effect ranged from 0.16 to 2.11 after household factors and travel time, and from 0.18 to 2.00 after also adjusting for the neighbouring private share.*

### S9 Table. Probability classes by division at thresholds 0.8 and 0.2, and 0.95 and 0.05, 2025

| Division | Clusters | Above national, unadjusted | Below national, unadjusted | Above 80%, unadjusted | Above, adjusted for household factors and travel time | Below, adjusted | Above, also adjusted for neighbouring private share | Below, also adjusted | Caesarean among facility births (%) | Private share of facility births (%) | Caesarean, private births (%, 95% CI) | Caesarean, public births (%, 95% CI) |
| --- | --- | --- | --- | --- | --- | --- | --- | --- | --- | --- | --- | --- |
| Barishal | 235 | 153 | 54 | 0 | 148 | 53 | 137 | 54 | 75.4 | 73.6 | 85.3 (81.1 to 89.5) | 42.7 (33.2 to 52.2) |
| Chattogram | 481 | 77 | 354 | 0 | 39 | 384 | 22 | 391 | 55.2 | 62.0 | 73.5 (70.1 to 76.9) | 22.9 (17.9 to 27.8) |
| Dhaka | 620 | 598 | 6 | 209 | 498 | 3 | 489 | 2 | 81.3 | 79.7 | 89.4 (87.4 to 91.4) | 46.0 (39.1 to 52.9) |
| Khulna | 418 | 418 | 0 | 287 | 418 | 0 | 418 | 0 | 83.6 | 82.0 | 92.4 (90.4 to 94.4) | 46.0 (37.9 to 54.2) |
| Mymensingh | 156 | 19 | 10 | 0 | 119 | 4 | 124 | 3 | 72.5 | 69.1 | 91.2 (86.7 to 95.7) | 29.6 (19.0 to 40.2) |
| Rajshahi | 335 | 187 | 58 | 18 | 173 | 83 | 174 | 73 | 74.5 | 74.2 | 89.0 (85.7 to 92.2) | 34.3 (27.1 to 41.4) |
| Rangpur | 315 | 51 | 49 | 0 | 54 | 17 | 65 | 5 | 73.6 | 69.7 | 91.4 (88.8 to 94.0) | 34.3 (26.6 to 42.0) |
| Sylhet | 160 | 0 | 148 | 0 | 0 | 157 | 0 | 157 | 60.6 | 62.8 | 82.0 (76.6 to 87.3) | 23.8 (16.0 to 31.7) |

*Counts of cluster locations where the smoothed surface has probability above 0.8 (above) or below 0.2 (below). These describe the model surface at cluster points; they are not independent cluster-level estimates. Cluster rates and shares are unweighted; sector rates are survey-weighted.*

**Stricter thresholds: cluster locations by posterior probability**

| Division | Clusters | Unadjusted: above 0.8 | Above 0.95 | Below 0.2 | Below 0.05 | Fully adjusted: above 0.8 | Above 0.95 | Below 0.2 | Below 0.05 |
| --- | --- | --- | --- | --- | --- | --- | --- | --- | --- |
| Barishal | 235 | 153 | 113 | 54 | 45 | 137 | 101 | 54 | 49 |
| Chattogram | 481 | 77 | 50 | 354 | 348 | 22 | 0 | 391 | 370 |
| Dhaka | 620 | 598 | 584 | 6 | 1 | 489 | 338 | 2 | 1 |
| Khulna | 418 | 418 | 416 | 0 | 0 | 418 | 415 | 0 | 0 |
| Mymensingh | 156 | 19 | 6 | 10 | 4 | 124 | 27 | 3 | 2 |
| Rajshahi | 335 | 187 | 141 | 58 | 23 | 174 | 131 | 73 | 10 |
| Rangpur | 315 | 51 | 0 | 48 | 2 | 65 | 9 | 5 | 0 |
| Sylhet | 160 | 0 | 0 | 148 | 127 | 0 | 0 | 157 | 142 |

*Unadjusted: probability that the smoothed caesarean rate among facility births exceeds the national rate. Fully adjusted: probability that the spatial effect is above zero after household factors, travel time and the neighbouring private share. Probabilities are conditional on the model.*

**Cluster summaries by probability class, unadjusted surface**

| Probability class | Clusters | Facility births | Caesarean rate (%) | Own-cluster private share (%) | Nearby private share (%) | Mean wealth quintile | Urban (%) |
| --- | --- | --- | --- | --- | --- | --- | --- |
| Below 0.2 | 679 | 1,729 | 51.6 | 58.2 | 58.5 | 3.27 | 31.9 |
| Uncertain | 538 | 1,275 | 73.6 | 70.8 | 70.9 | 2.77 | 23.8 |
| Above 0.8 | 1,503 | 3,671 | 81.9 | 80.2 | 79.5 | 3.21 | 26.5 |

*Probability that the smoothed caesarean rate among facility births exceeds the national rate of 72.8%. Unweighted cluster values; the nearby private share averages the eight nearest clusters. Mean wealth quintile runs from 1 (poorest) to 5 (richest).*

### S1 Fig. Directed acyclic graph of selection into facility and sector of birth

Solid arrows show paths through measured factors; dashed arrows show paths through unmeasured factors. Household wealth, education and residence, and modelled travel time with local supply, affect whether a woman gives birth in a facility and in which sector. Unmeasured obstetric indication affects facility birth, sector and caesarean; a caesarean planned before labour affects sector choice and caesarean. Analyses within sectors condition on facility birth (red box), which is affected by both measured and unmeasured factors. Among facility births, wealth and travel time can therefore become linked to unmeasured indication, which can bias comparisons of wealth or travel time within sectors. A direct path from wealth to caesarean (demand) is omitted for clarity.

### S2 Fig. Probability that the caesarean rate among facility births exceeded 80%, 2025

Binomial generalized additive model with a two-dimensional spatial smooth and 1,000 draws from the approximate posterior. Probabilities are conditional on the model. Solid lines mark a probability of 0.8 and dashed lines 0.2. Cluster coordinates are displaced for confidentiality. Boundaries: geoBoundaries.

### S3 Fig. Adjusted prevalence ratio by modelled travel time to the nearest health facility, restricted cubic splines, 2025

Survey-weighted Poisson models with a restricted cubic spline of modelled motorised travel time (three knots at 2.7, 12.2 and 27.3 minutes, the 10th, 50th and 90th percentiles), adjusted for age group, parity, wealth quintile, residence and education. Curves show the prevalence ratio against 5 minutes (dotted line) with 95% confidence bands, between the 5th and 95th percentiles of travel time (1.6 to 34.6 minutes); the rug marks cluster values. P values test the non-linear spline term (Wald test). Terracotta: private-sector births; navy: public-sector births. Post hoc analysis.
